# Assessing the Causal Effect of Blood Pressure on Renal Cancer in the UK Biobank via Mendelian Randomisation

**DOI:** 10.1101/2022.11.25.22282712

**Authors:** Lei Clifton, Xiaonan Liu, Jennifer A Collister, Thomas J Littlejohns, David J Hunter

## Abstract

1.

**Background:** Hypertension is associated with increasing risk of renal cancer in observational studies, but there is insufficient evidence on whether hypertension is a causal factor for renal cancer.

**Methods:** We analysed data from 313,520 individuals of White British ancestry aged 40-69 years at baseline from the UK Biobank prospective cohort. We used polygenic risk scores (PRS) for SBP and DBP, respectively, in Mendelian randomisation analyses to assess the causal effect of blood pressure on renal cancer. Our primary outcome is renal-parenchyma cancer, and we used bladder cancer as a negative control. We conducted additional sensitivity analyses, using individual SNPs from the PRS as instruments, to ensure the MR results are robust.

**Results:** Among the study population, 1159 participants developed renal-parenchyma cancer over the median of 12.5 years of follow-up. Every 5 mmHg increase in measured SBP and DBP was significantly associated with increased risk for renal-parenchyma cancer (HR: 1.04; 95% CI: 1.02-1.05; p < 0.001 and HR: 1.07; 95% CI: 1.03-1.10; p < 0.001 respectively). Consistent with observational association analyses, we observed statistically significant associations between each 5 mmHg genetically elevated BP and risk of renal-parenchyma cancer, with DBP (HR: 1.17; 95% CI: 1.07-1.27; p < 0.001) having a stronger association than SBP (HR: 1.09; 95% CI: 1.02-1.17; p = 0.009). These significant associations were maintained in sensitivity analyses. We observed a null association of genetically predicted blood pressure with bladder cancer.

**Conclusions:** These findings provide evidence that elevated BP is causally associated with renal-parenchyma cancer.

## 2. Introduction

Prior studies suggest a positive association between hypertension and renal cancer (Flaherty *et al*., 2005; Vatten *et al*., 2007; Weikert *et al*., 2008; Christakoudi *et al*., 2020), but it is not clear whether high blood pressure (BP) is a causal factor for renal cancer, as hypertension may be confounded by other risk factors such as obesity. Blood pressure (BP) has not been associated with renal-pelvis carcinoma (Chow *et al*., 2000), although such an association has been suggested (Liaw *et al*., 1997).

Evidence from prior studies is inconsistent on whether it is systolic blood pressure (SBP) or diastolic blood pressure (DBP), or both, that is associated with renal cancer. Some found that high DBP (but not SBP) increases the risk of renal cell carcinoma (RCC) (Johansson *et al*., 2019), while others reported an increased risk of RCC for both SBP and DBP among men (Chow *et al*., 2000; Häggström *et al*., 2013).

Mendelian randomisation (MR) is an application of the instrumental variable (IV) method for causal inference, where the genetic variation among people is utilised as the IV. If the genetic risk of disease is independent of environmental risk factors, these studies can be viewed as equivalent to randomised control trials (RCTs) conducted by nature that allows causal inference from observational studies under certain assumptions (Figure 1). The genetic instruments in earlier MR analyses were typically individual genetic variants, but they were susceptible to weak instrument bias. In recent years, large-scale genome-wide association (GWAS) studies have facilitated the discovery of a substantial number of genetic variants associated with risk factors and health conditions (Palmer *et al*., 2012; Burgess, Foley, *et al*., 2020). These genetic effects can be aggregated into a single score, often referred to as a polygenic risk score (PRS). Due to its simplicity, increased power and avoidance of weak instrument bias, the use of PRS as the genetic instrument in MR analyses is increasing (Burgess and Simon G Thompson, 2013; Holmes *et al*., 2015; Zekavat *et al*., 2021). The corresponding developments in MR methodology have allowed rigorous analyses and interpretation (Burgess, Davey Smith, *et al*., 2020; Burgess, Foley, *et al*., 2020).

**Figure 1:**
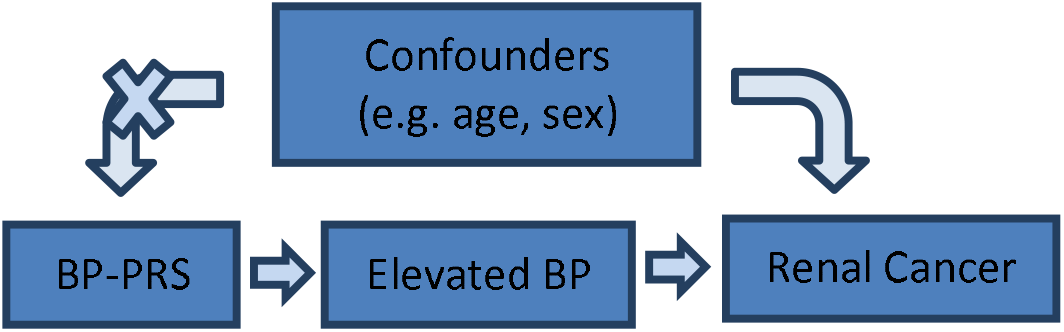
Mendelian randomisation permits causal inference between elevated BP and renal cancer, because the genetic instrument BP-PRS is not affected by confounders such as age and sex. BP: blood pressure, PRS: polygenic risk score.

In this study, we conducted MR using PRS for blood pressure (SBP-PRS and DBP-PRS, respectively) as genetic instruments to investigate whether blood pressure is a causal risk factor for renal cancer. We examined the key assumptions of MR, adopted methods for minimising potential biases, and conducted sensitivity analyses to establish the robustness of our MR results.

## 3. Methods

### 3.1 Study population and outcome definition

We analysed data from the UK Biobank (UKB), an ongoing large-scale population-based prospective cohort study of approximately 500,000 individuals recruited across the United Kingdom between March 2006 and October 2010 (UK Biobank Coordinating Centre, 2007; Collins, 2012). Our study population consists of participants who met UKB internal genetic quality control, were of White British ancestry, were aged 40-69 years old, and who had non-missing core variables (including measured SBP, DBP, Body mass index (BMI), and smoking status) at recruitment. Those with implausible BP values (SBP <70 mmHg or >270 mmHg; DBP <50 mmHg or >150 mmHg) at recruitment were excluded. We also excluded prevalent cases of cancer diagnosis (UKB Data Fields 40013 and 40006), except non-melanoma skin cancer (ICD-9: 173.X and ICD-10: C44.X) and carcinoma in situ (CIS) (ICD-9: 230.X-234.X and ICD-10: D00.X-D09.X).

For participants who were on antihypertensive medications (Tapela *et al*., 2021) at baseline assessment, we added 15 and 10 mmHg to their SBP and DBP measures, respectively (Warren *et al*., 2017; Zekavat *et al*., 2021).

Our primary outcome is renal-parenchyma cancer. Incident renal-parenchyma cancer cases were identified using International Classification of Diseases (ICD-9: 189.0 and ICD-10: C64.X) from (i) cancer registry data augmented with hospital inpatient data, and (ii) death registry data. The follow-up time for each participant was calculated as the number of years from the date of baseline assessment to the earliest of the following: date of renal-parenchyma diagnosis, date of death that is not caused by renal-parenchyma cancer, loss to follow-up date, or UKB administrative censoring date (England: 30 Sep 2021; Scotland: 31 Jul 2021; Wales: 31 Jul 2019).

The secondary outcome is the composite of both renal-parenchyma (ICD codes above) and renal-pelvis cancers (ICD-9: 189.1 and ICD-10: C65.X); individuals are regarded as experiencing the renal-composite outcome if they were diagnosed with either renal-parenchyma or renal-pelvis cancer. Censoring for the secondary outcome was defined analogously by replacing renal-parenchyma cancer with renal-composite cancer from above.

Our exposures of interest were SBP and DBP, for which we used SBP-PRS and DBP-PRS (described in Section 3.2) as the corresponding instrumental variables. Our primary analyses therefore consist of two parallel sets of analysis for SBP-PRS and DBP-PRS, respectively.

### 3.2 Calculating PRS

We identified suitable PRS from the literature, selecting scores comprised of BP-associated SNPs, with 884 SNPs in the SBP-PRS and 885 in the DBP-PRS (Evangelou *et al*., 2018). Effect sizes (i.e. *β*_*i*_ in the PRS equation below) for both SBP and DBP were derived from the International Consortium of Blood Pressure-Genome Wide Association Studies (ICBP), Million Veteran Program (MVP) and Estonian Genomic Centre of the University of Tartu (EGCUT).

We used the imputed genotyping data (version 3) from the UKB cohort; full details of the genotyping and imputation are described elsewhere (Bycroft *et al*., 2018). We computed the PRS using a published pipeline (Collister, Liu and Clifton, 2022), performing quality control for both SNPs and samples.

During SNP QC, we removed the following (Supplementary Table 1):

- SNPs with low imputation quality in UKB (imputation information score < 0.4),
- ambiguous SNPs (A/T or C/G SNPs with MAF > 0.49),
- rare variants with MAF < 0.005, and
- correlated variants, defined by linkage disequilibrium (LD) r^2^ ≥ 0.3 (Supplementary Table 1).

After applying SNP QC, we were left with 880 SBP-SNPs and 881 DBP-SNPs.

During sample QC, we excluded participants who were sex-discordant, outliers for missingness or heterozygosity, or related at 3^rd^ degree or higher, using UKB Data Field 22020 (Bycroft *et al*., 2018).

We then calculated the PRS of an individual *j* from the defined study population by the weighted sum of trait-associated SNPs,

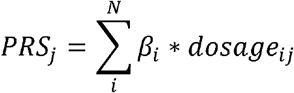

where N is the total number of SNPs, *β*_*i*_ is the effect size (or beta) of SNP *i*, and *dosage*_*ij*_ is the number of effect alleles (usually encoded as 0, 1 or 2 in SNP *i* for individual *j* for the effect allele).

### 3.3 Observational association of baseline blood pressure with incident renal cancer

Prior to MR analyses, we constructed Cox proportional hazard models to assess the association of baseline measured BP on incident renal cancer, adjusting for the following variables: age, sex, BMI, smoking status, weekly alcohol intake, weekly physical activity, SBP at baseline (and DBP at baseline, in a separate parallel model), number of comorbidities, Townsend deprivation index, household income, education, and UK country of residence. The proportional hazard assumption was assessed by visually examining the scaled Schoenfeld residuals.

### 3.4 MR methodology

In our main MR analysis, we performed MR analysis associating SBP-PRS and DBP-PRS, respectively, with renal cancer in the UKB. All genetic variants included in the PRS were assumed to be valid instruments in our main MR analysis (Gajendragadkar *et al*., 2021). Both the PRS-exposure and PRS-outcome associations used the same dataset (i.e. UKB), rendering our main analysis one-sample MR.

This study is reported in accordance with the Strengthening the Reporting of Observational Studies in Epidemiology - Mendelian Randomisation (STROBE-MR) guidelines (Smith *et al*., 2019; Burgess, Davey Smith, *et al*., 2020).

We used the triangulation approach to estimate the causal effect of baseline BP (i.e. exposure *X*) on renal cancer (i.e. outcome *Y*) via the genetic instrument BP-PRS (i.e. instrument *Z*). The ratio estimate or Wald estimate of the causal effect is (Teumer, 2018)

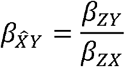

where *β*_*ZY*_ is the direct effect of the instrument on the outcome *Y, β*_*ZX*_ is the effect of the instrument on the exposure *X*, and 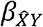 is the estimated causal effect of *X* on *Y*. The causal path way is *Z* via *X* to *Y*.

To minimise potential biases, we restricted the covariates in the models to the essential ones (Burgess, Davey Smith, *et al*., 2020), including age, sex, body mass index (BMI), smoking status, genotyping array, and the first 10 principal components (PCs). Association analysis of BP-PRS with incident renal cancer, *β*_*ZY*_, was performed using multivariable Cox proportional hazard model while the association between BP-PRS and BP, *β*_*ZX*_, was obtained by a linear regression adjusting for the same covariates; the estimated *β*_*ZX*_ also provided the check for the relevance assumption described below. We first used SBP-PRS in our models, and then repeated the analyses replacing SBP-PRS with DBP-PRS. Using PRS, an aggregated genetic score of multiple SNPs, as the genetic instrument in our main MR analysis, we were able to utilise the individual level data in the UKB to estimate both *β*_*ZY*_ and *β*_*ZX*_ in the above.

To assess the robustness of our MR results and elucidate the impacts of individual variants, we further conducted sensitivity analyses, where we considered the individual BP-SNPs as instruments in two-sample MR. In our two-sample MR sensitivity analyses, the instrument-exposure association, *β*_*ZX*_, were obtained from public GWAS summary statistics among cohorts external to the UKB, and the instrument-outcome association, *β*_*ZY*_ were estimated using the individual-level UKB data.

Two-sample MR uses two different study samples to estimate the instrument-exposure and instrument-outcome associations, respectively, where the two samples should have no overlap to avoid bias towards exposure-outcome association (Benn and Børge G Nordestgaard, 2018; Davies, Holmes and Davey Smith, 2018). Analogously, it is important to avoid sample overlap between the data in which the PRS was developed, and the data in which the PRS is used in analyses. Our study population from UKB is external to the development of the BP-PRS (Evangelou *et al*., 2018), hence is suitable for assessing the instrument-outcome association in the two-sample MR sensitivity analyses. The summary level associations during the development stage of the BP-PRS provides us with the instrument-exposure association in the two-sample MR as our sensitivity analyses.

Causal inferences via MR are subject to assumptions about the instrumental variables in order for them to be considered valid instruments. The three key assumptions of the genetic instrument are that it is strongly associated with the exposure of interest (relevance assumption), not related to any confounder of the exposure-outcome association (independence assumption), and affects the outcome only through the exposure of interest (exclusion restriction)(Davies, Holmes and Davey Smith, 2018). We checked each assumption as follows:

- Relevance assumption: The estimated *β*_*ZX*_ in the triangulation approach above checked for this assumption, where we fitted linear models regressing SBP-PRS on SBP, and DBP-PRS on DBP, respectively. The commonly used threshold of F-statistic > 10 indicates low risk of weak-instrument bias (Lawlor *et al*., 2008; Minelli *et al*., 2018; Zekavat *et al*., 2021).
- Independence assumption: The independence assumption is analogous to the balance of the baseline characteristics in an RCT; therefore we first examined such balance using the baseline table, and then conducted additional MR sensitivity analysis using PhenoScanner (Staley *et al*., 2016; Kamat *et al*., 2019) to identify SNPs associated with confounders.
- Exclusion restriction: the undesirable “horizontal pleiotropy” violates this assumption. We deployed bladder cancer (ICD-9: 188.X and ICD-10: C67.X) as our “negative control” outcome. We performed inverse variance weighted (IVW) heterogeneity test and MR-Egger intercept test to assess the existence of horizontal pleiotropy. Additionally, we conducted MR sensitivity analyses, including IVW multiplicative random-effect method (Burgess, Butterworth and Thompson, 2013), MR-Egger method (Bowden, Smith and Burgess, 2015) and weighted median-based method (Bowden *et al*., 2016) that relax this assumption.

### 3.5 Sensitivity analyses

In our analysis, since our instrument is a PRS constructed from multiple genetic variants, there is the additional assumption that all SNPs in our PRS are also valid instruments (Pierce, Ahsan and Vanderweele, 2011; Burgess and Simon G. Thompson, 2013). Since both BMI and smoking are known confounders that influence both BP and renal cancer (Chow *et al*., 2000; Christakoudi *et al*., 2020), we performed sensitivity analysis using the PRS after removing confounder-associated SNPs.

For each BP-SNP, we used the PhenoScanner (Staley *et al*., 2016; Kamat *et al*., 2019) search tool to identify any published associations (p-value < 5e-8) with any of the following renal cancer-associated traits: BMI (<u>trait ID</u>: <u>EFO_0004340</u>), obesity (<u>EFO_0001073</u>) and smoking status (<u>EFO_0006527</u>). The tool also simultaneously searches for association between proxies (defined as SNPs that are in high LD, r^2^ > 0.8 with the BP-SNP) and the specified traits. In situation where only a proxy was found to be associated with the trait, the corresponding BP-SNP was regarded as associated with the trait as well. We repeated the process with a more conservative SNP-trait association p-value threshold, 5e-6, to further test the independence assumption.

There is no known association between BP and bladder cancer, hence we chose it to be our “negative control” outcome for examining potential pleiotropic effects (Burgess, Davey Smith, *et al*., 2020). If our genetic instrument affects renal cancer solely through blood pressure, then it should not have any effect on bladder cancer. We fitted a Cox regression model with bladder cancer as the outcome, adjusting for the same covariates as in the main MR analysis where SBP-PRS and DBP-PRS were the genetic instruments, respectively.

Two-sample MR was further conducted to assess the robustness of our main MR results. We first computed the association coefficient between each SNP in the PRS and incident renal-parenchyma cancer (i.e. *β*_*ZY*_) in our UKB dataset by fitting a logistic regression for each SNP, adjusting for age, sex, BMI, smoking status, genotyping array and first 10 PCs. The instrument-exposure association for each SNP (i.e. *β*_*ZX*_) is the effect size given for the PRS developed in Evangelou *et al*., derived in cohorts external to the UKB, rendering these analyses two-sample MR (Benn and Børge G. Nordestgaard, 2018).

In our IVW estimate sensitivity analysis, a ratio estimate (i.e. Wald estimate) was computed for each SNP, and then combined into one causal estimate. To account for the heterogeneity due to pleiotropic effects, we used multiplicative random-effects models instead of fixed-effects ones (Burgess, Davey Smith, *et al*., 2020). The MR-Egger method replaces the exclusion restriction assumption with a weaker assumption called Instrument Strength Independent of Direct Effect (InSIDE) assumption, where it assumes pleiotropic effects of the SNPs on the outcome are uncorrelated with *β*_*ZX*_. The modelling of MR-Egger allows for both direct pleiotropic effects and indirect causal effects, but its InSIDE assumption is difficult to check, and its causal estimate can be sensitive to outlying variants. Therefore, we further performed the weighted median method which has a natural robustness to outlying variants (Burgess, Davey Smith, *et al*., 2020). The weighted median method uses inverse variance weights, assuming at least half of the genetic variants are valid instruments.

In addition, we performed leave-one-out analyses with the IVW method, to further investigate the influence of outlying variants. To examine the impact of potential hidden population structure, we removed the first 10 PCs in a sensitivity analysis. Finally, the Steiger directionality test (Hemani, Tilling and Davey Smith, 2017) was conducted to test whether the direction of causality (i.e. our exposure causes the outcome) is valid.

In a Mendelian Randomization study, higher diastolic blood pressure (DBP), but not systolic blood pressure (SBP), was found to increase the risk of renal cell carcinoma (Johansson *et al*., 2019). To assess the independent causal effect of SBP and DBP, we conducted a sensitivity MR analysis that includes both SBP and DBP in the model.

To explore whether the association between BP and renal cancer is different for men and for women, we conducted a sensitivity analysis by analysing men and women in separate models (i.e. stratified analysis by sex).

Statistical significance was declared at a two-tailed 5% significance level. PRS calculation and logistic regressions for two-sample MR were conducted using PLINK 2.0; all remaining analyses were performed in R version 4.0.2. IVW heterogeneity test, IVW method, MR-Egger method, and weighted median-based method were performed using the “Mendelian Randomization” package in R. Steiger directionality test and leave-one-out analyses were performed using the “TwoSampleMR” package in R.

## 4. Results

### 4.1 Baseline characteristics

A total of N = 313,520 genotyped individuals met the inclusion criteria and passed genetic QC (Figure 2). Among these, 1159 developed renal-parenchyma cancer (median follow-up = 12.5 years), and 89 developed renal-pelvis cancer (median follow-up = 12.5 years). 22 individuals developed cancer of both the renal-parenchyma and renal-pelvis (Supplementary Figure 1).

**Figure 2:**
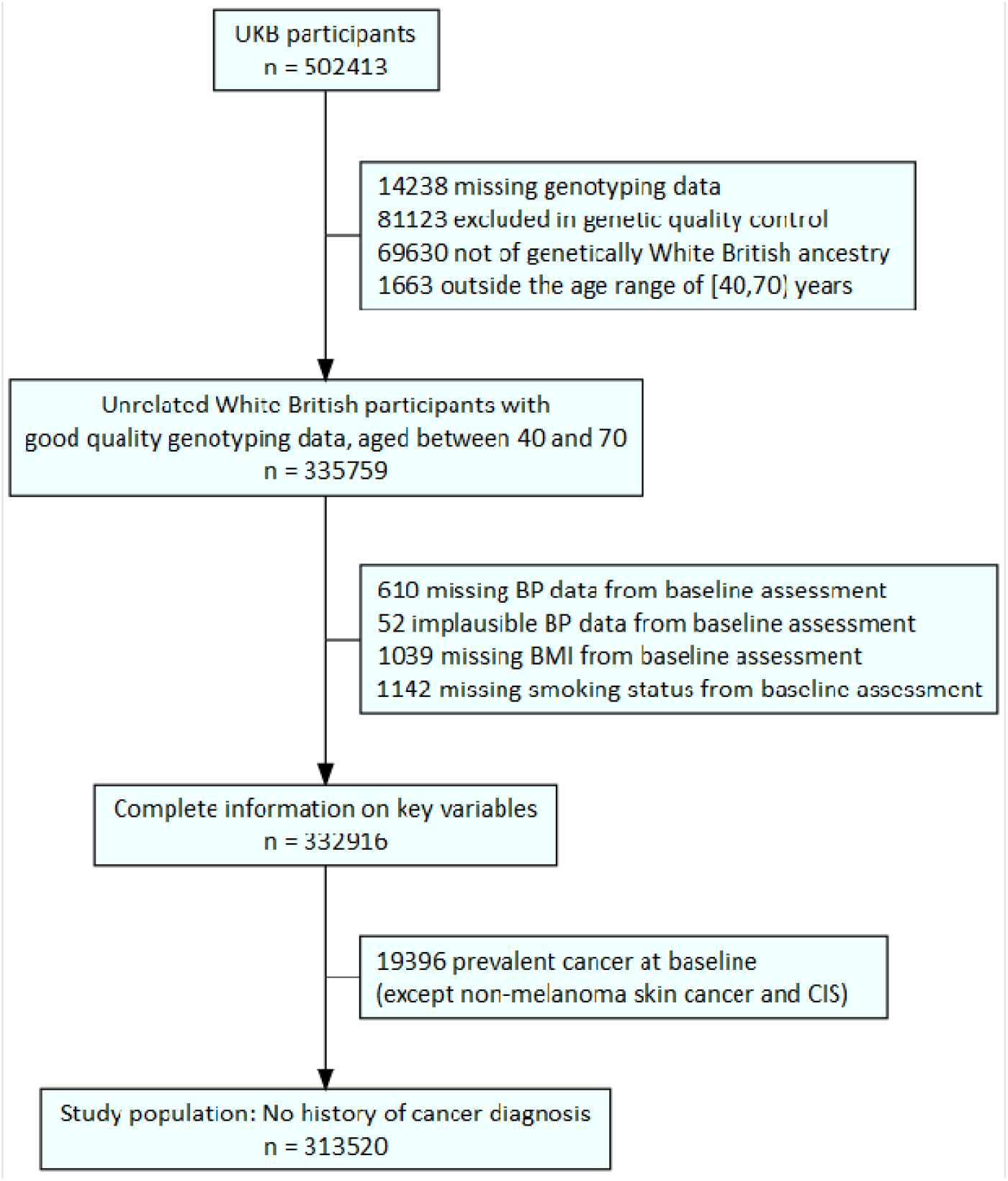
Flowchart illustrating study population

The characteristics of our study population are presented in Table 1 (detailed baseline characteristics in Supplementary Tables 2-3).

**Table 1.**
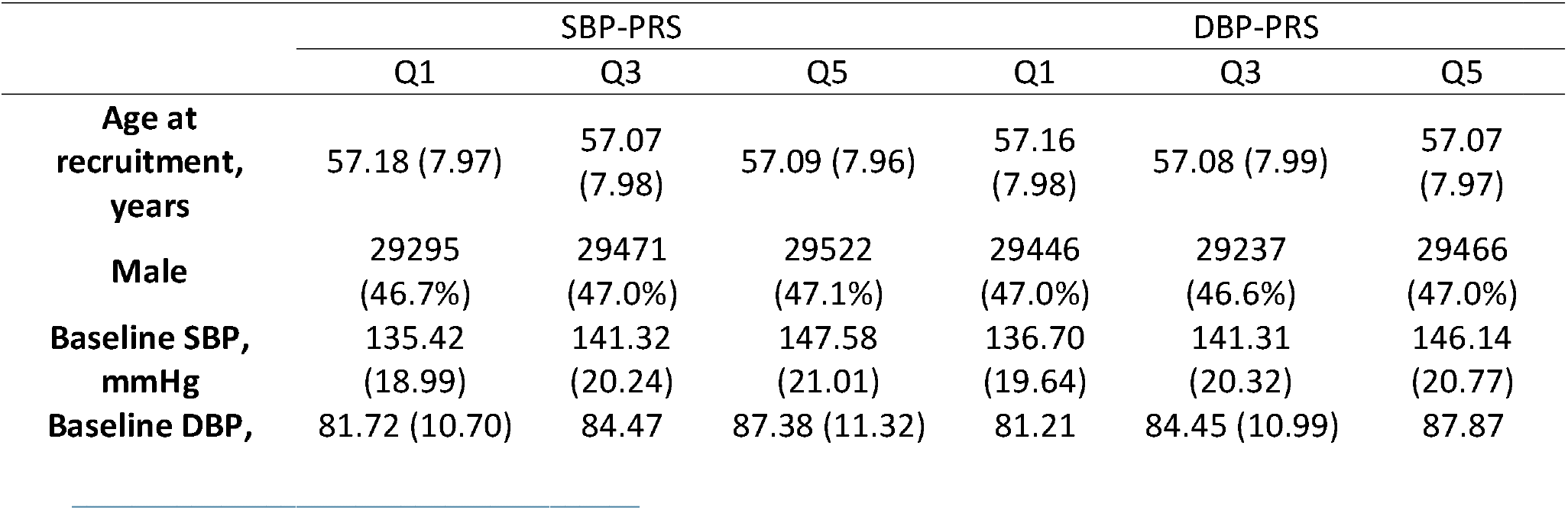
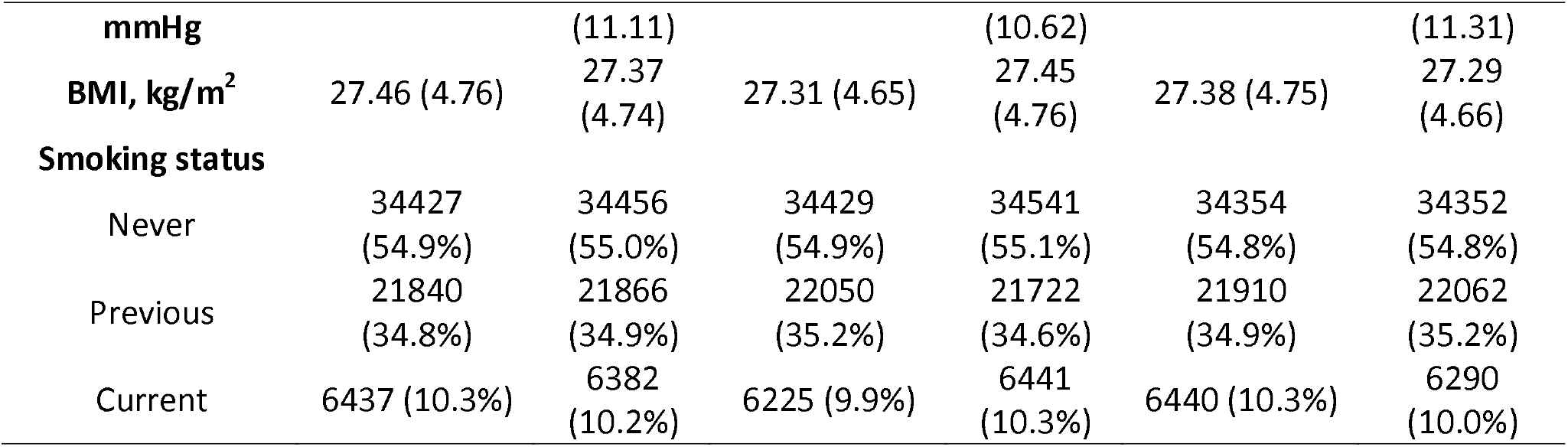
Baseline characteristics of the study population by strata of SBP- and DBP-PRS (N = 313,520 in total, n = 62,704 in each quintile). Mean (SD) are presented for continuous variables, frequency (percentage) are reported for categorical variables. Percentages may not add up to 100 due to rounding. PRS: Polygenic risk scores. SBP: Systolic blood pressure. DBP: Diastolic blood pressure. Q1: the 1^st^ (lowest) quintile. Q3: the 3^rd^ (middle) quintile. Q5: the 5^th^ (highest) quintile.

### 4.2 Observational association of baseline BP with incident renal cancer

Figure 3 shows that both baseline SBP and DBP were significantly associated with risk of renal-parenchyma cancer, after adjustment for covariates described in the Methods section. The hazard ratio (HR) per 5 mmHg increase in SBP was 1.04 (95% confidence interval [CI]: 1.02 - 1.05, p < 0.001); the HR per 5 mmHg increase in DBP was 1.07 (95% CI: 1.03-1.10, p < 0.001). The full output of the Cox model are shown in Supplementary Table 4. No violations of proportional hazard assumptions were observed. We observed similar results for the secondary outcome (renal-composite cancer, Supplementary Table 5 and Supplementary Figure 5).

**Figure 3:**
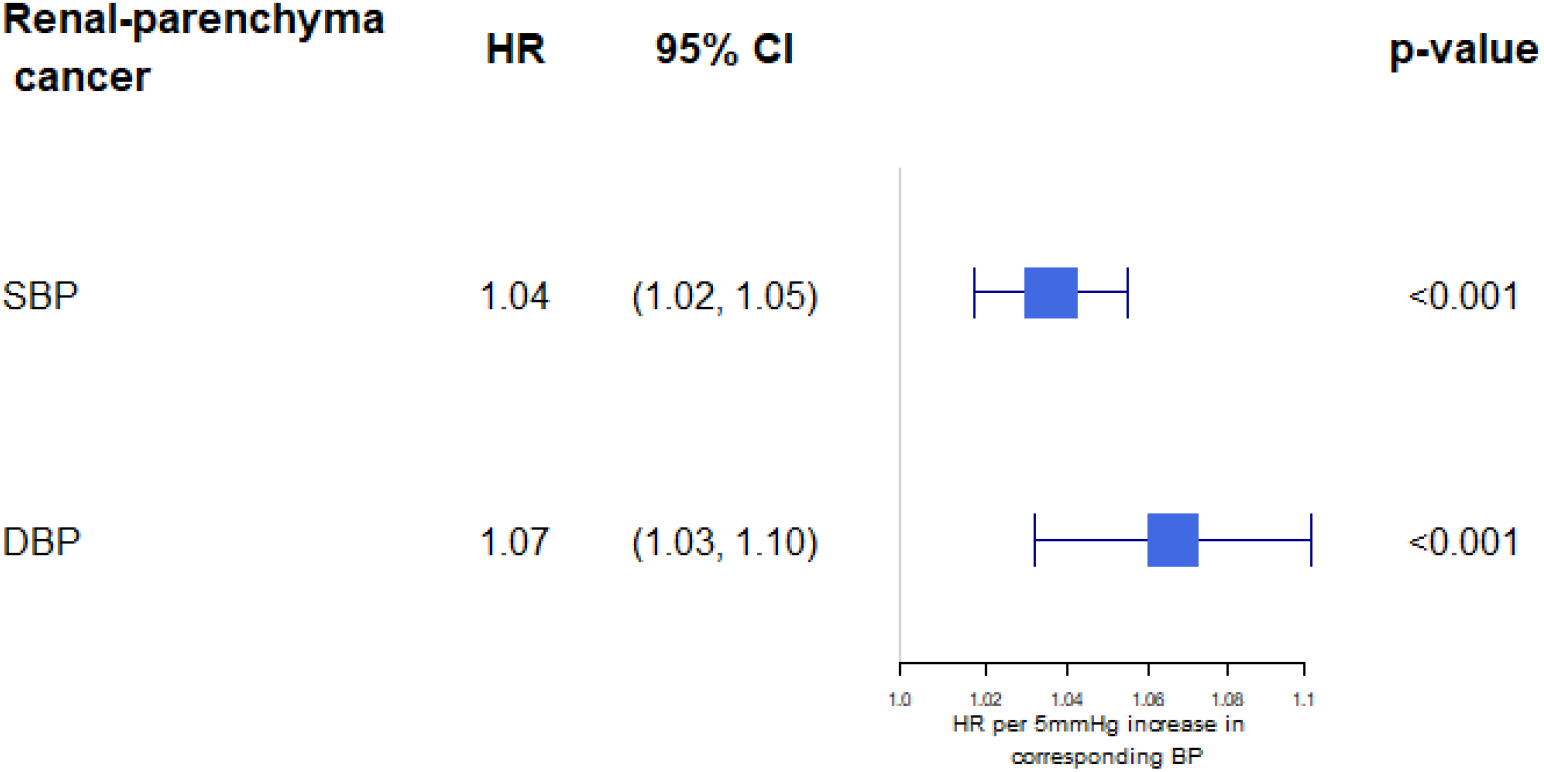
Observational association of BP with renal-parenchyma cancer via a multivariable Cox model adjusted for age, sex, BMI, smoking status, weekly alcohol intake, weekly physical activity, the number of comorbidities, Townsend deprivation index, household income, education, and UK country of residence.

**Figure 4.**
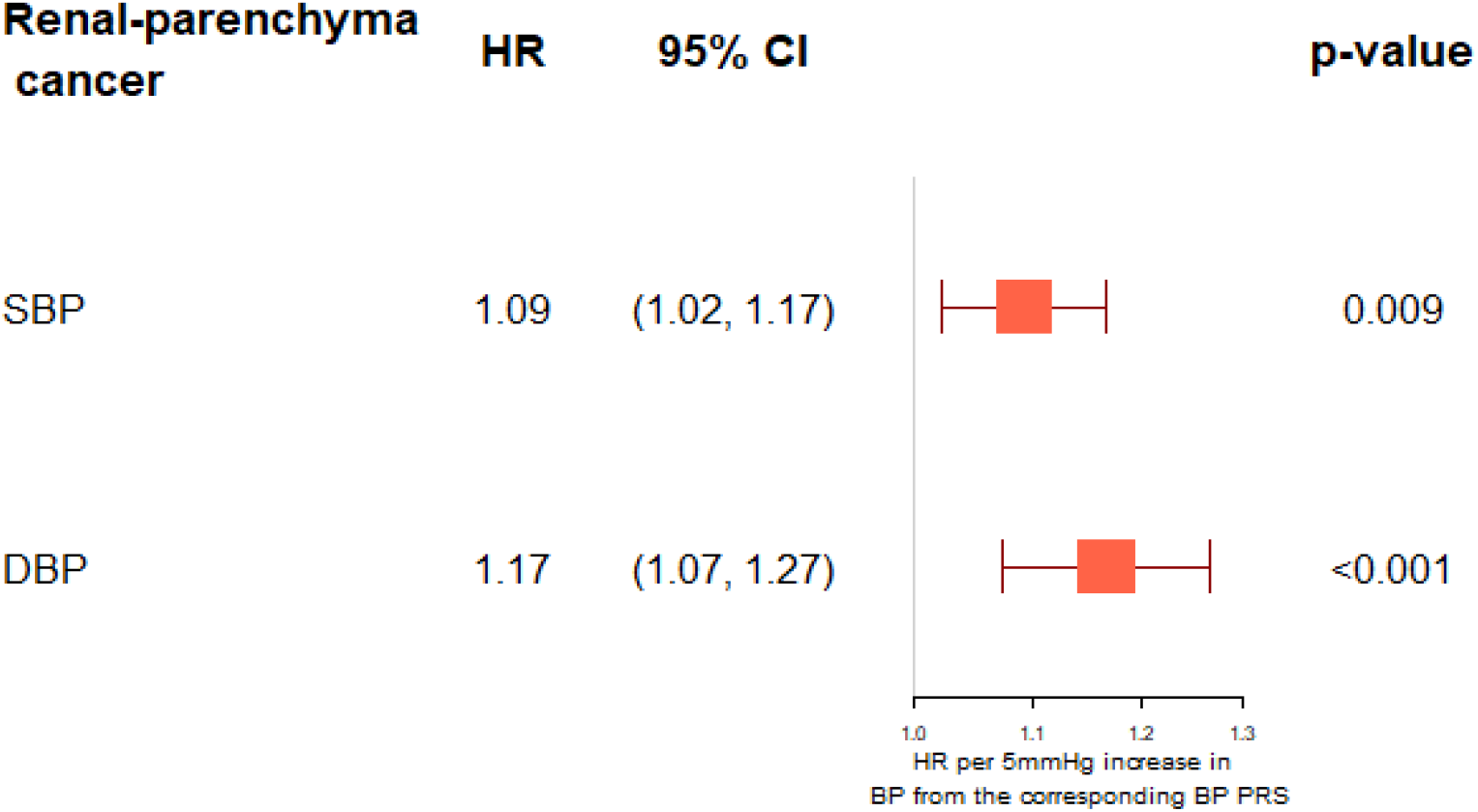
Genetic association of BP conferred by the corresponding BP-PRS with renal-parenchyma cancer. All models were adjusted for age, sex, BMI, smoking status, genotype array and first 10 genetic PCs.

### 4.3 Genetic association of baseline BP with incident renal cancer

Our SBP-PRS and DBP-PRS consisted of 880 and 881 SNPs after QC, respectively. **Error! Reference source not found**. shows the genetic association of renal-parenchyma cancer obtained from our main one-sample MR analysis. Each 5 mmHg increase in SBP conferred by SBP-PRS was significantly associated with the risk of renal-parenchyma cancer (HR: 1.09; 95% CI: 1.02-1.17; p = 0.01). Similarly, every 5 mmHg increase in DBP conferred by DBP-PRS was also significantly associated with the risk of renal-parenchyma cancer (HR: 1.17; 95% CI: 1.07-1.27; p < 0.001). DBP showed stronger effect than SBP on renal-parenchyma cancer, according to the two parallel models described in Methods. Similar results were observed for the secondary outcome (renal-composite cancer, Supplementary Figure 6).

### 4.4 Sensitivity analyses

The estimated *β*_*ZX*_ from the triangulation approach was not only part of the causal estimation but also provided checks for the relevance assumption (Supplementary Table 6). For each mmHg increase of SBP-PRS (F-statistic = 6,239), SBP is increased by 1.151 mmHg. For each mmHg increase of DBP-PRS (F-statistic = 4,634), DBP is increased by 1.056 mmHg. These high F-statistics are well above the common threshold of 10, indicating that both SBP-PRS and DBP-PRS are strong instrument variables.

To check the independence assumption, we chose two p-value thresholds, 5e-8 (i.e. variant-trait association p-value < 5e-8) and 5e-6, to identify SNPs significantly associated with BMI, obesity or smoking status. Using p-value threshold 5e-8, we removed total of 43 SNPs from BP-PRS, including 37 SNPs associated with BMI (<u>trait ID</u>: EFO_0004340), 3 SNPs associated with obesity (EFO_0001073) as defined by the original GWAS, and 3 SNPs associated with smoking status (EFO_0006527) were removed from BP-PRS. With a p-value threshold of 5e-6, a total of 71 SNPs (58 associated with BMI, 6 associated with obesity, and 7 associated with smoking status) were removed from BP-PRS. Both the SBP- and DBP-PRS still satisfied the relevance assumption after removing confounder-associated SNPs (Supplementary Tables 7-8). We observed that causal estimates of both SBP and DBP were attenuated after removing confounder associated SNPs (Supplementary Figure 2). The attenuation of the causal estimate appeared more distinct on the SBP than DBP.

Our results using bladder cancer as the negative control outcome support the absence of horizontal pleiotropy, shown in Supplementary Tables 9-10. The HR of bladder cancer is 0.97 (95% CI: 0.91-1.03, p = 0.35) per 5 mmHg increase in SBP conveyed by SBP-PRS, and is 1.03 (95% CI: 0.92-1.16, p = 0.61) per 5 mmHg increase in DBP conveyed by DBP-PRS.

Across SBP and DBP genetic instruments, the IVW heterogeneity test (p = 0.55 for SBP and p = 0.59 for DBP) and the MR-Egger intercept term (p = 0.47 for SBP and p = 0.88 for DBP) were both insignificant, suggesting negligible contribution of horizontal pleiotropy (Supplementary Table 11).

Two-sample Mendelian randomization was additionally performed as sensitivity analyses, where we used the publicly available association estimates of SNPs with BP (derived *external to* the UKB), and derived the association estimates of those SNPs with renal-parenchyma cancer (derived *within* the UKB). The IVW multiplicative random effect yielded comparable estimates (OR: 1.11; 95% CI: 1.03-1.20; p = 0.01 and OR: 1.27; 95% CI: 1.12-1.44; p < 0.001 for SBP and DBP respectively) to the main analyses as anticipated. The MR-Egger method and weighted median method also showed similarly significant results (Supplementary Table 11 and Supplementary Figures 3-4). In particular, MR-Egger method and weighted median method showed a wider 95% confidence interval compared with our main MR analysis. DBP retained its more pronounced effect than SBP on renal-parenchyma cancer.

Leave-one-out analyses with the IVW method (Supplementary materials in pdf) showed similar causal estimates across BP genetic variants, indicating no particular SNP drives the observed association. The Steiger directionality test suggest the correct temporal direction of BP causing renal-parenchyma cancer. There is no major difference in the estimates after removing the first 10 PCs (Supplementary Tables 12-13), indicating the absence of confounding by ancestry, hence supporting the similarity of the two samples in our two-sample MR method.

To explore the independent causal effect of SBP and DBP, we repeated our main MR analyses but further adjusted for SBP-PRS when estimating the association of DBP-PRS with renal-parenchyma cancer (i.e. both SBP-PRS and DBP-PRS are included in the model). The effect of DBP remained significant with renal-parenchyma cancer (HR per 5 mmHg increase from DBP-PRS: 1.31; 95% CI: 1.07-1.61; p = 0.01) whereas the effect of SBP was no longer statistically significant (HR per 5 mmHg increase from SBP-PRS: 0.97; 95% CI: 0.86-1.08; p = 0.55). This indicates a potential dependency between SBP and DBP in contributing to renal-parenchyma cancer, where DBP (instead of SBP) may be the actual causal factor of renal-parenchyma cancer. We note the relatively high correlation between SBP and DBP (*ρ* = 0.8).

Stratified analyses by sex in two separate models indicated a stronger relationship between DBP and renal-parenchyma cancer among males than among females; the pattern of association for SBP was similar but less pronounced (Supplementary Table 14). To further investigate the potential difference between men and women, we added an interaction term SBP×sex in the above MR model for SBP. Similarly, we added an interaction term DBP×sex in the above MR model for DBP. We did not observe statistically significant interactions for renal-parenchyma cancer (p_SBP×sex_ = 0.88, p_DBP×sex_ = 0.61). Similar results were observed for renal-composite cancer (Supplementary Table 15).

## 5. Discussion

These findings may have important implications for the prevention of renal cancer. We found that both higher SBP and DBP were associated with renal-parenchyma cancer in traditional epidemiological analyses. Our MR analyses support that elevated DBP is causally linked to increased risk for developing renal-parenchyma cancer.

A major strength of this study is the large sample size in combination with detailed data collection, including genotyping and longitudinal linkage to electronic medical records over a long follow-up period. In addition, our study design involves a combination of observational association analyses and Mendelian randomisation, which provide a full scope of robust investigation from association to causal inference.

One limitation of one-sample MR is its reduced statistical power compared with two-sample MR, and therefore we performed two-sample MR sensitivity analyses to ensure the robustness of our results. However, the two separate samples in two-sample MR may not come from the same underlying populations, causing potential violation of the instrument variable assumption. In our two-sample MR sensitivity analyses, this problem is largely mitigated by the fact that (i) both the derivation population of our BP-PRS and our defined study population within UKB are of European origin, and (ii) we further adjusted for the first 10 PCs of ancestry in our analyses. However, there is still the possibility that the two samples may differ on finer strata of ethnic groups or population characteristics. Furthermore, our findings are only applicable to individuals of white British ancestry, and further studies are needed to validate our findings in other ancestries.

Another limitation of our study is that as the number of SNPs included in PRS increased, the genetic variants are more often associated with multiple traits, and therefore we conducted sensitivity analyses by excluding SNPs associated with potential confounders (e.g. BMI). Our MR sensitivity analysis showed that the causal link between blood pressure and renal cancer stands after removing confounder-associated SNPs from our BP-PRS. We have kept such exclusions to the minimum during the sensitivity analyses, but their results may still be potentially biased.

Our findings were similar to those from the European Prospective Investigation into Cancer and Nutrition (EPIC) study (Alcala *et al*., 2022) that also observed a causal relationship of DBP in renal cancer. The authors used both the EPIC cohort containing 715 incident renal cell carcinoma (RCC) cases (N = 278,309), and the UKB cohort containing 977 incident RCC cases (N = 422,718). However, our approaches are distinct from and complementary to those in the EPIC study in the following aspects: (i) we used PRS in our main MR analysis, and then conducted two-sample MR as sensitivity analyses, whereas Alcala *et al*. only did the latter; (ii) our GWAS summary statistics are derived from a population external to UKB, whereas Alcala *et al*. identified BP genetic variants (i.e. variant-exposure) by conducting a GWAS study using the UKB data, and then obtained variant-outcome estimates from public RCC GWAS summary statistics (Scelo *et al*., 2017); (iii) we used bladder cancer as our negative control outcome, and demonstrated the absence of horizontal pleiotropy; (iv) we conducted stratified MR analysis by sex, and investigated whether genetically predicted BP affects men differently from women in risks of developing renal cancers.

Both ureter and bladder cancers are distinct from, but related to, renal cancer. Neither is related to BP, but we chose bladder (not ureter) cancer as our negative control outcome to check the independence assumption and the exclusion restriction assumption (Davies, Holmes and Davey Smith, 2018), because bladder cancer is more common than ureter cancer in UKB and ureter cancer is more likely to be anatomically involved with renal cancer.

Through observational and genetic association analyses, we showed that elevated DBP is a causal factor for renal cancer. The mechanisms linking BP to renal cancer are not yet well established, but it has been suggested that angiogenic and other growth factors may be involved in renal carcinogenesis (Chow *et al*., 2000). Our results suggest that maintaining adequate control of BP could be one approach for preventing renal cancer.

## Supporting information

MR leave-one-out sensitivity analysis for DBP on outcome

MR leave-one-out sensitivity analysis for SBP on outcome

## Data Availability

This research has been conducted using the UK Biobank Resource under Application Number 33952. Requests to access the data should be made via application directly to the UK Biobank, https://www.ukbiobank.ac.uk

https://github.com/xiaonanl1996/MRforRenal

## 6. Declarations

### 6.1 Ethics approval and consent to participate

The UK Biobank study received ethical approval from the North West Multi-centre Research Ethics Committee (REC reference: 11/NW/03820). All participants gave written informed consent before enrolment in the study, which was conducted in accordance with the principles of the Declaration of Helsinki. This study has been conducted using the UK Biobank Resource under Application Number 33952.

### 6.2 Patient and community involvement

The analyses presented here are based on existing data from the UK Biobank cohort study, and the authors were not involved in participant recruitment. To the best of our knowledge, no patients were explicitly engaged in the design or implementation of the UK Biobank study. No patients were asked to advise on interpretation or writing these results. Results from UK Biobank are routinely disseminated to study participants via the study website and social media outlets.

### 6.3 Consent for publication

Yes.

### 6.4 Availability of data and material

This research has been conducted using the UK Biobank Resource under Application Number 33952. Requests to access the data should be made via application directly to the UK Biobank, *https://www.ukbiobank.ac.uk*

The code used for analyses are available at *https://github.com/xiaonanl1996/MRforRenal*

### 6.7 Author contributions

DH conceived the project; LC, XL, and DH outlined the statistical methods; LC and XL drafted the manuscripts; XL conducted the statistical analysis; JC reviewed the R scripts by XL; TL reviewed the manuscript. All authors have revised the manuscript and agreed on its contents.

### 6.8 Transparency statement

The lead author affirms that this manuscript is an honest, accurate, and transparent account of the study being reported; that no important aspects of the study have been omitted; and that any discrepancies from the study as planned have been explained.

## 6.9 Acknowledgements

We thank the participants and staff of the UK Biobank for enabling us to conduct this research.

## References

Alcala, K. et al. (2022) ‘The relationship between blood pressure and risk of renal cell carcinoma’, International Journal of Epidemiology. Oxford University Press (OUP), 2022, pp. 1–11. doi: 10.1093/IJE/DYAC042.

Benn, M. and Nordestgaard Børge G (2018) ‘From genome-wide association studies to Mendelian randomization: Novel opportunities for understanding cardiovascular disease causality, pathogenesis, prevention, and treatment’, Cardiovascular Research. Oxford Academic, pp. 1192–1208. doi: 10.1093/cvr/cvy045.

Benn, M. and Nordestgaard, Børge G. (2018) ‘From genome-wide association studies to Mendelian randomization: Novel opportunities for understanding cardiovascular disease causality, pathogenesis, prevention, and treatment’, Cardiovascular Research. Oxford Academic, pp. 1192–1208. doi: 10.1093/cvr/cvy045.

Bowden, J. et al. (2016) ‘Consistent Estimation in Mendelian Randomization with Some Invalid Instruments Using a Weighted Median Estimator’, Genetic epidemiology. Genet Epidemiol, 40(4), pp. 304–314. doi: 10.1002/GEPI.21965.

Bowden, J., Smith, G. D. and Burgess, S. (2015) ‘Mendelian randomization with invalid instruments: effect estimation and bias detection through Egger regression’, International journal of epidemiology. Int J Epidemiol, 44(2), pp. 512–525. doi: 10.1093/IJE/DYV080.

Burgess, S., Foley, C. N., et al. (2020) ‘A robust and efficient method for Mendelian randomization with hundreds of genetic variants’, Nature Communications 2020 11:1. Nature Publishing Group, 11(1), pp. 1–11. doi: 10.1038/s41467-019-14156-4.

Burgess, S., Davey Smith, G., et al. (2020) ‘Guidelines for performing Mendelian randomization investigations’, Wellcome Open Research. F1000 Research Limited, 4, p. 186. doi: 10.12688/wellcomeopenres.15555.2.

Burgess, S., Butterworth, A. and Thompson, S. G. (2013) ‘Mendelian randomization analysis with multiple genetic variants using summarized data’, Genetic epidemiology. Genet Epidemiol, 37(7), pp. 658–665. doi: 10.1002/GEPI.21758.

Burgess, S. and Thompson Simon G (2013) ‘Use of allele scores as instrumental variables for Mendelian randomization’, International Journal of Epidemiology. Oxford Academic, 42(4), pp. 1134–1144. doi: 10.1093/IJE/DYT093.

Burgess, S. and Thompson, Simon G. (2013) ‘Use of allele scores as instrumental variables for Mendelian randomization’, International Journal of Epidemiology, 42(4), pp. 1134–1144. doi: 10.1093/IJE/DYT093.

Bycroft, C. et al. (2018) ‘The UK Biobank resource with deep phenotyping and genomic data’, Nature, 562(7726), pp. 203–209. doi: 10.1038/s41586-018-0579-z.

Chow, W.-H. et al. (2000) ‘Obesity, Hypertension, and the Risk of Kidney Cancer in Men’, New England Journal of Medicine. Massachusetts Medical Society, 343(18), pp. 1305–1311. doi: 10.1056/nejm200011023431804.

Christakoudi, S. et al. (2020) ‘Blood pressure and risk of cancer in the European Prospective Investigation into Cancer and Nutrition’, International Journal of Cancer. Int J Cancer, 146(10), pp. 2680–2693. doi: 10.1002/ijc.32576.

Collins, R. (2012) ‘What makes UK Biobank special?’, The Lancet, pp. 1173–1174. doi: 10.1016/S0140-6736(12)60404-8.

Collister, J. A., Liu, X. and Clifton, L. (2022) ‘Calculating Polygenic Risk Scores (PRS) in UK Biobank: A Practical Guide for Epidemiologists’, Frontiers in Genetics. Frontiers, 0, p. 105. doi: 10.3389/FGENE.2022.818574.

Davies, N. M., Holmes, M. V. and Davey Smith, G. (2018) ‘Reading Mendelian randomisation studies: A guide, glossary, and checklist for clinicians’, BMJ (Online), 362. doi: 10.1136/bmj.k601.

Evangelou, E. et al. (2018) ‘Genetic analysis of over 1 million people identifies 535 new loci associated with blood pressure traits’, Nature Genetics. Nature Publishing Group, 50(10), pp. 1412–1425. doi: 10.1038/s41588-018-0205-x.

Flaherty, K. T. et al. (2005) ‘A prospective study of body mass index, hypertension, and smoking and the risk of renal cell carcinoma (United States)’, Cancer Causes and Control. Springer, 16(9), pp. 1099–1106. doi: 10.1007/s10552-005-0349-8.

Gajendragadkar, P. R. et al. (2021) ‘Assessment of the causal relevance of ECG parameters for risk of atrial fibrillation: A mendelian randomisation study’, PLoS Medicine. Public Library of Science, 18(5), p. e1003572. doi: 10.1371/journal.pmed.1003572.

Häggström, C. et al. (2013) ‘Metabolic Factors Associated with Risk of Renal Cell Carcinoma’, PLOS ONE. Public Library of Science, 8(2), p. e57475. doi: 10.1371/JOURNAL.PONE.0057475.

Hemani, G., Tilling, K. and Davey Smith, G. (2017) ‘Orienting the causal relationship between imprecisely measured traits using GWAS summary data’, PLOS Genetics. Public Library of Science, 13(11), p. e1007081. doi: 10.1371/JOURNAL.PGEN.1007081.

Holmes, M. V. et al. (2015) ‘Mendelian randomization of blood lipids for coronary heart disease’, European Heart Journal. Eur Heart J, 36(9), pp. 539–550. doi: 10.1093/eurheartj/eht571.

Johansson, M. et al. (2019) ‘The influence of obesity-related factors in the etiology of renal cell carcinoma-A mendelian randomization study’, PLoS Medicine, 16(1), pp. 1–16. doi: 10.1371/journal.pmed.1002724.

Kamat, M. A. et al. (2019) ‘PhenoScanner V2: an expanded tool for searching human genotype-phenotype associations’, Bioinformatics (Oxford, England). Bioinformatics, 35(22), pp. 4851–4853. doi: 10.1093/BIOINFORMATICS/BTZ469.

Lawlor, D. A. et al. (2008) ‘Mendelian randomization: Using genes as instruments for making causal inferences in epidemiology’, Statistics in Medicine. John Wiley & Sons, Ltd, 27(8), pp. 1133–1163. Available at: https://onlinelibrary.wiley.com/doi/full/10.1002/sim.3034 (Accessed: 13 March 2020).

Liaw, K. L. et al. (1997) ‘Possible relation between hypertension and cancers of the renal pelvis and ureter’, International Journal of Cancer. Wiley-Liss, Inc.* Received, 70(3), pp. 265–268. doi: 10.1002/(SICI)1097-0215(19970127)70:3<265::AID-IJC3>3.0.CO;2-V.

Minelli, C. et al. (2018) ‘Age at puberty and risk of asthma: A Mendelian randomisation study’, PLOS Medicine. Public Library of Science, 15(8), p. e1002634. doi: 10.1371/JOURNAL.PMED.1002634.

Palmer, T. M. et al. (2012) ‘Using multiple genetic variants as instrumental variables for modifiable risk factors’, in Statistical Methods in Medical Research, pp. 223–242. doi: 10.1177/0962280210394459.

Pierce, B. L., Ahsan, H. and Vanderweele, T. J. (2011) ‘Power and instrument strength requirements for Mendelian randomization studies using multiple genetic variants’, International Journal of Epidemiology. Oxford Academic, 40(3), pp. 740–752. doi: 10.1093/IJE/DYQ151.

Scelo, G. et al. (2017) ‘Genome-wide association study identifies multiple risk loci for renal cell carcinoma’, Nature Communications, 8, pp. 1–9. doi: 10.1038/ncomms15724.

Smith, G. D. et al. (2019) ‘STROBE-MR: Guidelines for strengthening the reporting of Mendelian randomization studies’. PeerJ Inc. doi: 10.7287/PEERJ.PREPRINTS.27857V1.

Staley, J. R. et al. (2016) ‘PhenoScanner: a database of human genotype-phenotype associations’, Bioinformatics (Oxford, England). Bioinformatics, 32(20), pp. 3207–3209. doi: 10.1093/BIOINFORMATICS/BTW373.

Tapela, N. et al. (2021) ‘Prevalence and determinants of hypertension control among almost 100 000 treated adults in the UK’, BMJ Open Heart. Archives of Disease in childhood, 8(1), p. e001461. doi: 10.1136/openhrt-2020-001461.

Teumer, A. (2018) ‘Common Methods for Performing Mendelian Randomization’, Frontiers in Cardiovascular Medicine. Frontiers, 0, p. 51. doi: 10.3389/FCVM.2018.00051.

UK Biobank Coordinating Centre (2007) ‘UK Biobank: Protocol for a large-scale prospective epidemiological resource UK Biobank Coordinating Centre Stockport’, UKBB-PROT-09-06 (Main Phase), 06(March), pp. 1–112. doi: 10.1126/science.311.5767.1535c.

Vatten, L. J. et al. (2007) ‘Blood pressure and renal cancer risk: The HUNT Study in Norway’, British Journal of Cancer, 97(1), pp. 112–114. doi: 10.1038/SJ.BJC.6603823.

Warren, H. R. et al. (2017) ‘Genome-wide association analysis identifies novel blood pressure loci and offers biological insights into cardiovascular risk’, Nature Genetics. Nature Research, 49(3), pp. 403–415. doi: 10.1038/ng.3768.

Weikert, S. et al. (2008) ‘Blood pressure and risk of renal cell carcinoma in the European prospective investigation into cancer and nutrition’, American Journal of Epidemiology, 167(4), pp. 438–446. doi: 10.1093/aje/kwm321.

Zekavat, S. M. et al. (2021) ‘Elevated Blood Pressure Increases Pneumonia Risk: Epidemiological Association and Mendelian Randomization in the UK Biobank’, Med. Cell Press, 2(2), pp. 137-148.e4. doi: 10.1016/J.MEDJ.2020.11.001.

