## Supplementary figures and images for "Assessing the Causal Effect of Blood Pressure on Renal Cancer in the UK Biobank via Mendelian Randomisation"

### MR leave-one-out sensitivity analysis for DBP on outcome

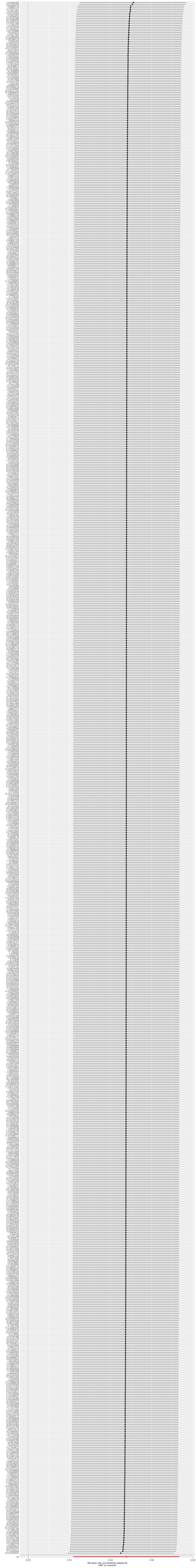

### MR leave-one-out sensitivity analysis for SBP on outcome

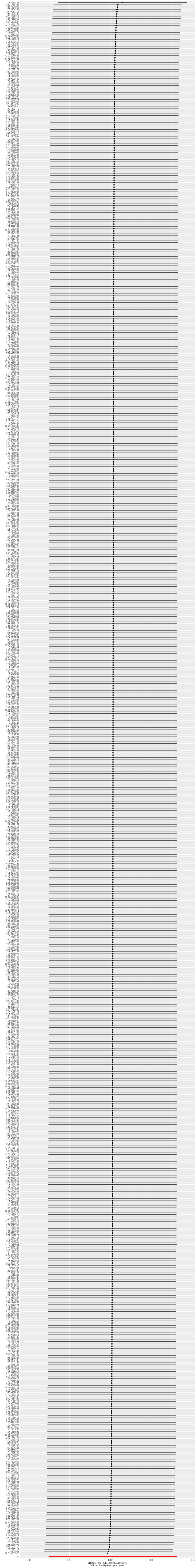
