## Supplementary Tables and Figures for "Assessing the Causal Effect of Blood Pressure on Renal Cancer in the UK Biobank via Mendelian Randomisation"

Lei Clifton^1^, Xiaonan Liu^1^, Jennifer A Collister^1^, Thomas J Littlejohns^1^, David J Hunter^1,2^

1. Nuffield Department of Population Health, University of Oxford, UK.

2. Department of Epidemiology, Harvard TH Chan School of Public Health, Boston, USA

### 1 Table of SNP QC

Supplementary Table 1: SNP QC. All the 884 SNPs in the SBP-PRS are present in the DBP-PRS, but the effect size of each SNP is different in the SBP-PRS and DBP-PRS. PRS: Polygenic risk scores. nSNPs: number of SNPs included in the PRS. QC: Quality control. UKB: UK Biobank. MAF: Minor allele frequency. LD: Linkage disequilibrium.

| Disease | PRS | nSNPs | Unavailable in UKB | Imputation info < 0.4 | Ambiguous | MAF $<$0.005 | LD  r^2^ ≥ 0.3 | Remaining SNPs |
| --- | --- | --- | --- | --- | --- | --- | --- | --- |
| Hypertension | SBP | 884 | 0 | 0 | 3 | 0 | 1 | 880 |
|  | DBP | 885 | 0 | 0 | 3 | 0 | 1 | 881 |

### 2 Renal-parenchyma and renal-pelvis cancers

n=22

Incident renal-parenchyma cancer n=1137

Incident renal-pelvis cancer n=67

Supplementary Figure 1: Venn diagram showing the overlap between incident renal-parenchyma and renal-pelvis cancers. A total of 1159 individuals developed renal-parenchyma cancer, among whom 22 also had renal-pelvis cancer. Note that the size of diagram does not represent the actual size.

### 3 Baseline characteristics

#### 3.1 Baseline characteristics by PRS

Supplementary Table 2: Detailed baseline characteristics of the study population by strata of SBP and DBP-PRS (N = 313,520 in total, n=62,704 in each quintile). Mean (SD) are presented for continuous variables, frequency (percentage) are reported for categorical variables. Percentages may not add up to 100 due to rounding. SBP: Systolic blood pressure. DBP: Diastolic blood pressure. PRS: Polygenic risk scores. BMI: Body Mass Index.

|  | SBP-PRS | | | DBP-PRS | | |
| --- | --- | --- | --- | --- | --- | --- |
|  | Q1: lowest | Q3 | Q5: highest | Q1: lowest | Q3 | Q5: highest |
| **Age at recruitment, years** | 57.18 (7.97) | 57.07 (7.98) | 57.09 (7.96) | 57.16 (7.98) | 57.08 (7.99) | 57.07 (7.97) |
| **Male** | 29295 (46.7%) | 29471 (47.0%) | 29522 (47.1%) | 29446 (47.0%) | 29237 (46.6%) | 29466 (47.0%) |
| **Baseline SBP, mmHg** | 135.42 (18.99) | 141.32 (20.24) | 147.58 (21.01) | 136.70 (19.64) | 141.31 (20.32) | 146.14 (20.77) |
| **Baseline DBP, mmHg** | 81.72 (10.70) | 84.47 (11.11) | 87.38 (11.32) | 81.21 (10.62) | 84.45 (10.99) | 87.87 (11.31) |
| **BMI, kg/m^2^** | 27.46 (4.76) | 27.37 (4.74) | 27.31 (4.65) | 27.45 (4.76) | 27.38 (4.75) | 27.29 (4.66) |
| **Smoking status** |  |  |  |  |  |  |
| Never | 34427 (54.9%) | 34456 (55.0%) | 34429 (54.9%) | 34541 (55.1%) | 34354 (54.8%) | 34352 (54.8%) |
| Previous | 21840 (34.8%) | 21866 (34.9%) | 22050 (35.2%) | 21722 (34.6%) | 21910 (34.9%) | 22062 (35.2%) |
| Current | 6437 (10.3%) | 6382 (10.2%) | 6225 (9.9%) | 6441 (10.3%) | 6440 (10.3%) | 6290 (10.0%) |
| **Alcohol units per week** |  |  |  |  |  |  |
| None reported | 10472 (16.7%) | 10334 (16.5%) | 10569 (16.9%) | 10487 (16.7%) | 10557 (16.8%) | 10575 (16.9%) |
| Less than 5 units | 12116 (19.3%) | 11931 (19.0%) | 11992 (19.1%) | 12088 (19.3%) | 11949 (19.1%) | 12174 (19.4%) |
| 5 to 10 units | 10396 (16.6%) | 10528 (16.8%) | 10386 (16.6%) | 10364 (16.5%) | 10356 (16.5%) | 10259 (16.4%) |
| 10 to 20 units | 14779 (23.6%) | 14907 (23.8%) | 14663 (23.4%) | 14793 (23.6%) | 15024 (24.0%) | 14719 (23.5%) |
| 20 to 30 units | 7426 (11.8%) | 7393 (11.8%) | 7566 (12.1%) | 7454 (11.9%) | 7335 (11.7%) | 7519 (12.0%) |
| 30 units or more | 7515 (12.0%) | 7611 (12.1%) | 7528 (12.0%) | 7518 (12.0%) | 7483 (11.9%) | 7458 (11.9%) |
| **Weekly physical activity, high (MET minutes > 1200)** | 32782 (64.2%) | 32742 (64.0%) | 32542 (64.0%) | 32765 (64.1%) | 32672 (64.1%) | 32690 (64.2%) |
| Missing | 11607 | 11580 | 11875 | 11605 | 11714 | 11802 |
| **Number of comorbidities** |  |  |  |  |  |  |
| 0 | 40737 (65.0%) | 40350 (64.3%) | 39604 (63.2%) | 40583 (64.7%) | 40235 (64.2%) | 39781 (63.4%) |
| 1 | 17123 (27.3%) | 17349 (27.7%) | 17659 (28.2%) | 17265 (27.5%) | 17412 (27.8%) | 17573 (28.0%) |
| 2 | 3964 (6.3%) | 4063 (6.5%) | 4378 (7.0%) | 3927 (6.3%) | 4132 (6.6%) | 4316 (6.9%) |
| >=3 | 880 (1.4%) | 942 (1.5%) | 1063 (1.7%) | 929 (1.5%) | 925 (1.5%) | 1034 (1.6%) |
| **Townsend Deprivation Index quintiles** |  |  |  |  |  |  |
| Q1: least deprived | 12531 (20.0%) | 12429 (19.8%) | 12613 (20.1%) | 12484 (19.9%) | 12494 (19.9%) | 12481 (19.9%) |
| Q2 | 12431 (19.8%) | 12506 (20.0%) | 12500 (20.0%) | 12424 (19.8%) | 12577 (20.1%) | 12595 (20.1%) |
| Q3 | 12585 (20.1%) | 12510 (20.0%) | 12658 (20.2%) | 12672 (20.2%) | 12346 (19.7%) | 12525 (20.0%) |
| Q4 | 12611 (20.1%) | 12687 (20.3%) | 12208 (19.5%) | 12525 (20.0%) | 12642 (20.2%) | 12356 (19.7%) |
| Q5: most deprived | 12468 (19.9%) | 12490 (19.9%) | 12654 (20.2%) | 12523 (20.0%) | 12572 (20.1%) | 12668 (20.2%) |
| Missing | 78 | 82 | 71 | 76 | 73 | 79 |
| **Household Income, GBP** |  |  |  |  |  |  |
| Less than 18,000 | 11488 (21.1%) | 11420 (21.0%) | 11771 (21.7%) | 11488 (21.1%) | 11551 (21.3%) | 11488 (21.2%) |
| 18,000 to 30,999 | 13773 (25.2%) | 13824 (25.5%) | 14040 (25.8%) | 13695 (25.2%) | 13653 (25.2%) | 13998 (25.8%) |
| 31,000 to 51,999 | 14471 (26.5%) | 14482 (26.7%) | 14453 (26.6%) | 14593 (26.8%) | 14434 (26.6%) | 14506 (26.8%) |
| 52,000 to 100,000 | 11738 (21.5%) | 11545 (21.3%) | 11272 (20.7%) | 11596 (21.3%) | 11526 (21.2%) | 11350 (20.9%) |
| Greater than 100,000 | 3086 (5.7%) | 3044 (5.6%) | 2796 (5.1%) | 3069 (5.6%) | 3114 (5.7%) | 2870 (5.3%) |
| Missing | 8148 | 8389 | 8372 | 8263 | 8426 | 8492 |
| **Highest level of education (ISCED)** |  |  |  |  |  |  |
| 5: Tertiary | 30299 (48.7%) | 30099 (48.4%) | 29498 (47.4%) | 30075 (48.4%) | 29993 (48.2%) | 29589 (47.6%) |
| 4: Post-secondary non-tertiary | 7726 (12.4%) | 7739 (12.4%) | 7621 (12.3%) | 7672 (12.3%) | 7663 (12.3%) | 7682 (12.3%) |
| 2-3: Secondary | 14048 (22.6%) | 14003 (22.5%) | 14496 (23.3%) | 14068 (22.6%) | 14103 (22.7%) | 14438 (23.2%) |
| 1: Primary | 10133 (16.3%) | 10384 (16.7%) | 10575 (17.0%) | 10382 (16.7%) | 10439 (16.8%) | 10502 (16.9%) |
| Missing | 498 | 479 | 514 | 507 | 506 | 493 |
| **UK country of residence** |  |  |  |  |  |  |
| England | 55148 (87.9%) | 55153 (88.0%) | 55526 (88.6%) | 54938 (87.6%) | 55287 (88.2%) | 55641 (88.7%) |
| Scotland | 4757 (7.6%) | 4758 (7.6%) | 4452 (7.1%) | 4910 (7.8%) | 4706 (7.5%) | 4365 (7.0%) |
| Wales | 2799 (4.5%) | 2793 (4.5%) | 2726 (4.3%) | 2856 (4.6%) | 2711 (4.3%) | 2698 (4.3%) |

#### 3.2 Baseline characteristics by sex

Supplementary Table 3: Detailed baseline characteristics of study population by sex (N=313,520). Mean (SD) are presented for continuous variables, frequency (percentage) are reported for categorical variables. Percentages may not add up to 100 due to rounding. SBP: Systolic blood pressure. DBP: Diastolic blood pressure. PRS: Polygenic risk scores. BMI: Body Mass Index.

|  | Female (N=166,794) | Male (N=146,726) | Total (N=313,520) |
| --- | --- | --- | --- |
| **Age at recruitment, years** | 56.90 (7.89) | 57.34 (8.06) | 57.10 (7.97) |
| **SBP-PRS** | -1.32 (3.81) | -1.31 (3.82) | -1.32 (3.82) |
| **DBP-PRS** | -2.98 (2.30) | -2.98 (2.30) | -2.98 (2.30) |
| **Baseline SBP, mmHg** | 138.14 (21.10) | 145.02 (19.30) | 141.36 (20.57) |
| **Baseline DBP, mmHg** | 82.48 (11.06) | 86.77 (10.98) | 84.49 (11.23) |
| **BMI, kg/m^2^** | 26.98 (5.10) | 27.82 (4.21) | 27.38 (4.72) |
| **Smoking status** |  |  |  |
| Never | 99640 (59.7%) | 72463 (49.4%) | 172103 (54.9%) |
| Previous | 52749 (31.6%) | 56797 (38.7%) | 109546 (34.9%) |
| Current | 14405 (8.6%) | 17466 (11.9%) | 31871 (10.2%) |
| **Alcohol units per week** |  |  |  |
| None reported | 35871 (21.5%) | 16548 (11.3%) | 52419 (16.7%) |
| Less than 5 units | 40318 (24.2%) | 20029 (13.7%) | 60347 (19.2%) |
| 5 to 10 units | 33050 (19.8%) | 18699 (12.7%) | 51749 (16.5%) |
| 10 to 20 units | 37033 (22.2%) | 37278 (25.4%) | 74311 (23.7%) |
| 20 to 30 units | 12878 (7.7%) | 24383 (16.6%) | 37261 (11.9%) |
| 30 units or more | 7644 (4.6%) | 29789 (20.3%) | 37433 (11.9%) |
| **Weekly physical activity, High (MET minutes > 1200)** | 82719 (63.5%) | 80746 (64.8%) | 163465 (64.1%) |
| Missing | 36555 | 22141 | 58696 |
| **Number of comorbidities** |  |  |  |
| 0 | 105744 (63.4%) | 95457 (65.1%) | 201201 (64.2%) |
| 1 | 46791 (28.1%) | 39987 (27.3%) | 86778 (27.7%) |
| 2 | 11493 (6.9%) | 9304 (6.3%) | 20797 (6.6%) |
| >=3 | 2766 (1.7%) | 1978 (1.3%) | 4744 (1.5%) |
| **Townsend Deprivation Index quintiles** |  |  |  |
| Q1: least deprived | 33153 (19.9%) | 29481 (20.1%) | 62634 (20.0%) |
| Q2 | 33452 (20.1%) | 29179 (19.9%) | 62631 (20.0%) |
| Q3 | 33581 (20.2%) | 29051 (19.8%) | 62632 (20.0%) |
| Q4 | 33878 (20.3%) | 28749 (19.6%) | 62627 (20.0%) |
| Q5: most deprived | 32540 (19.5%) | 30085 (20.5%) | 62625 (20.0%) |
| Missing | 190 | 181 | 371 |
| **Household Income, GBP** |  |  |  |
| Less than 18,000 | 32396 (23.3%) | 25048 (18.9%) | 57444 (21.2%) |
| 18,000 to 30,999 | 36697 (26.4%) | 32324 (24.4%) | 69021 (25.4%) |
| 31,000 to 51,999 | 36155 (26.0%) | 36442 (27.5%) | 72597 (26.7%) |
| 52,000 to 100,000 | 27097 (19.5%) | 30382 (23.0%) | 57479 (21.2%) |
| Greater than 100,000 | 6825 (4.9%) | 8081 (6.1%) | 14906 (5.5%) |
| Missing | 27624 | 14449 | 42073 |
| **Highest level of education (ISCED)** |  |  |  |
| 5: Tertiary | 71817 (43.4%) | 77920 (53.5%) | 149737 (48.1%) |
| 4: Post-secondary non-tertiary | 22638 (13.7%) | 15797 (10.9%) | 38435 (12.4%) |
| 2-3: Secondary | 43299 (26.2%) | 27493 (18.9%) | 70792 (22.8%) |
| 1: Primary | 27766 (16.8%) | 24309 (16.7%) | 52075 (16.7%) |
| Missing | 1274 | 1207 | 2481 |
| **UK country of residence** |  |  |  |
| England | 146668 (87.9%) | 129805 (88.5%) | 276473 (88.2%) |
| Scotland | 12765 (7.7%) | 10503 (7.2%) | 23268 (7.4%) |
| Wales | 7361 (4.4%) | 6418 (4.4%) | 13779 (4.4%) |

### 4 Observational association

Supplementary Table 4: Observational association of BP with incident renal-parenchyma cancer, N = 226,682. Models 1 and 2 assess the effect of baseline SBP and DBP, respectively, on renal-parenchyma cancer. HR of SBP and DBP represents the HR per 5mmHg increase in BP.

|  |  | Model 1 | | | Model 2 | | |
| --- | --- | --- | --- | --- | --- | --- | --- |
| Variable | Levels | HR | 95% CI | p | HR | 95% CI | p |
| Baseline SBP, mmHg |  | 1.04 | (1.02, 1.05) | <0.001 |  |  |  |
| Baseline DBP, mmHg |  |  |  |  | 1.07 | (1.03, 1.10) | <0.001 |
| Age at recruitment, years |  | 1.06 | (1.05, 1.07) | <0.001 | 1.06 | (1.05, 1.08) | <0.001 |
| Gender | Female | 1 |  |  | 1 |  |  |
|  | Male | 2.20 | (1.88, 2.58) | <0.001 | 2.18 | (1.86, 2.55) | <0.001 |
| BMI, kg/m^2^ |  | 1.04 | (1.03, 1.06) | <0.001 | 1.04 | (1.02, 1.05) | <0.001 |
| Smoking status | Never | 1 |  |  | 1 |  |  |
|  | Previous | 1.18 | (1.02, 1.38) | 0.030 | 1.19 | (1.02, 1.38) | 0.027 |
|  | Current | 1.63 | (1.30, 2.05) | <0.001 | 1.64 | (1.31, 2.05) | <0.001 |
| Alcohol units per week | None reported | 1 |  |  | 1 |  |  |
|  | Less than 5 units | 0.88 | (0.70, 1.10) | 0.250 | 0.88 | (0.70, 1.09) | 0.245 |
|  | 5 to 10 units | 0.76 | (0.59, 0.96) | 0.024 | 0.75 | (0.59, 0.96) | 0.021 |
|  | 10 to 20 units | 0.68 | (0.54, 0.85) | <0.001 | 0.68 | (0.54, 0.84) | <0.001 |
|  | 20 to 30 units | 0.48 | (0.36, 0.64) | <0.001 | 0.48 | (0.36, 0.64) | <0.001 |
|  | 30 units or more | 0.59 | (0.46, 0.76) | <0.001 | 0.59 | (0.46, 0.76) | <0.001 |
| Weekly physical activity | Low (MET minutes <= 1200) | 1 |  |  | 1 |  |  |
|  | High (MET minutes > 1200) | 0.85 | (0.73, 0.98) | 0.023 | 0.85 | (0.74, 0.98) | 0.029 |
| Number of comorbidities | 0 | 1 |  |  | 1 |  |  |
|  | 1 | 1.06 | (0.90, 1.24) | 0.474 | 1.06 | (0.91, 1.24) | 0.473 |
|  | 2 | 1.35 | (1.06, 1.71) | 0.015 | 1.35 | (1.07, 1.72) | 0.013 |
|  | >=3 | 1.35 | (0.86, 2.14) | 0.195 | 1.36 | (0.86, 2.15) | 0.185 |
| Townsend Deprivation Index quintiles | Q1: least deprived | 1 |  |  | 1 |  |  |
|  | Q2 | 0.85 | (0.68, 1.05) | 0.137 | 0.85 | (0.68, 1.05) | 0.135 |
|  | Q3 | 1.05 | (0.85, 1.29) | 0.648 | 1.05 | (0.85, 1.29) | 0.651 |
|  | Q4 | 0.87 | (0.70, 1.09) | 0.226 | 0.87 | (0.70, 1.08) | 0.215 |
|  | Q5: most deprived | 0.80 | (0.63, 1.00) | 0.053 | 0.79 | (0.63, 1.00) | 0.050 |
| Household Income, GBP | Less than 18,000 | 1 |  |  | 1 |  |  |
|  | 18,000 to 30,999 | 0.93 | (0.76, 1.12) | 0.430 | 0.93 | (0.76, 1.12) | 0.436 |
|  | 31,000 to 51,999 | 0.91 | (0.73, 1.12) | 0.376 | 0.91 | (0.73, 1.12) | 0.365 |
|  | 52,000 to 100,000 | 0.89 | (0.70, 1.14) | 0.371 | 0.89 | (0.70, 1.14) | 0.355 |
|  | Greater than 100,000 | 0.86 | (0.57, 1.28) | 0.453 | 0.85 | (0.57, 1.28) | 0.436 |
| Highest level of education (ISCED) | 5: Tertiary | 1 |  |  | 1 |  |  |
|  | 4: Post-secondary non-tertiary | 1.10 | (0.88, 1.37) | 0.391 | 1.10 | (0.88, 1.37) | 0.386 |
|  | 2-3: Secondary | 1.19 | (1.00, 1.43) | 0.056 | 1.20 | (1.00, 1.43) | 0.053 |
|  | 1: Primary | 1.10 | (0.90, 1.35) | 0.352 | 1.11 | (0.91, 1.36) | 0.312 |
| UK country of residence | England | 1 |  |  | 1 |  |  |
|  | Scotland | 0.95 | (0.73, 1.23) | 0.677 | 0.95 | (0.73, 1.23) | 0.686 |
|  | Wales | 0.58 | (0.37, 0.92) | 0.020 | 0.58 | (0.37, 0.91) | 0.019 |

Supplementary Table 5: Observational association of BP with incident renal-composite cancer N = 226,682. Models 1 and 2 assess the effect of baseline SBP and DBP, respectively, on renal-composite cancer. HR of SBP and DBP represents the HR per 5mmHg increase in BP.

|  |  | Model 1 | | | Model 2 | | |
| --- | --- | --- | --- | --- | --- | --- | --- |
| Variable | Levels | HR | 95% CI | p | HR | 95% CI | p |
| Baseline SBP, mmHg |  | 1.04 | (1.02, 1.05) | <0.001 |  |  |  |
| Baseline DBP, mmHg |  |  |  |  | 1.07 | (1.03, 1.10) | <0.001 |
| Age at recruitment, years |  | 1.06 | (1.05, 1.07) | <0.001 | 1.07 | (1.06, 1.08) | <0.001 |
| Gender | Female | 1 |  |  | 1 |  |  |
|  | Male | 2.20 | (1.89, 2.56) | <0.001 | 2.18 | (1.87, 2.54) | <0.001 |
| BMI, kg/m^2^ |  | 1.04 | (1.03, 1.05) | <0.001 | 1.04 | (1.02, 1.05) | <0.001 |
| Smoking status | Never | 1 |  |  | 1 |  |  |
|  | Previous | 1.19 | (1.03, 1.38) | 0.022 | 1.19 | (1.03, 1.39) | 0.019 |
|  | Current | 1.73 | (1.39, 2.14) | <0.001 | 1.73 | (1.39, 2.15) | <0.001 |
| Alcohol units per week | None reported | 1 |  |  | 1 |  |  |
|  | Less than 5 units | 0.88 | (0.71, 1.10) | 0.263 | 0.88 | (0.71, 1.10) | 0.258 |
|  | 5 to 10 units | 0.74 | (0.59, 0.94) | 0.014 | 0.74 | (0.58, 0.94) | 0.013 |
|  | 10 to 20 units | 0.70 | (0.56, 0.87) | 0.001 | 0.70 | (0.56, 0.86) | <0.001 |
|  | 20 to 30 units | 0.54 | (0.42, 0.71) | <0.001 | 0.54 | (0.42, 0.70) | <0.001 |
|  | 30 units or more | 0.59 | (0.46, 0.76) | <0.001 | 0.59 | (0.46, 0.76) | <0.001 |
| Weekly physical activity | Low (MET minutes <= 1200) | 1 |  |  | 1 |  |  |
|  | High (MET minutes > 1200) | 0.85 | (0.74, 0.98) | 0.023 | 0.86 | (0.75, 0.99) | 0.030 |
| Number of comorbidities | 0 | 1 |  |  | 1 |  |  |
|  | 1 | 1.07 | (0.92, 1.25) | 0.353 | 1.07 | (0.92, 1.25) | 0.351 |
|  | 2 | 1.31 | (1.03, 1.65) | 0.026 | 1.31 | (1.04, 1.66) | 0.023 |
|  | >=3 | 1.27 | (0.80, 2.00) | 0.309 | 1.28 | (0.81, 2.01) | 0.295 |
| Townsend Deprivation Index quintiles | Q1: least deprived | 1 |  |  | 1 |  |  |
|  | Q2 | 0.86 | (0.70, 1.06) | 0.163 | 0.86 | (0.70, 1.06) | 0.160 |
|  | Q3 | 1.04 | (0.85, 1.27) | 0.713 | 1.04 | (0.85, 1.27) | 0.716 |
|  | Q4 | 0.87 | (0.70, 1.07) | 0.192 | 0.86 | (0.70, 1.07) | 0.182 |
|  | Q5: most deprived | 0.80 | (0.64, 1.00) | 0.051 | 0.80 | (0.64, 1.00) | 0.048 |
| Household Income, GBP | Less than 18,000 | 1 |  |  | 1 |  |  |
|  | 18,000 to 30,999 | 0.90 | (0.75, 1.08) | 0.262 | 0.90 | (0.75, 1.08) | 0.266 |
|  | 31,000 to 51,999 | 0.93 | (0.76, 1.14) | 0.506 | 0.93 | (0.76, 1.14) | 0.494 |
|  | 52,000 to 100,000 | 0.90 | (0.71, 1.15) | 0.402 | 0.90 | (0.71, 1.14) | 0.384 |
|  | Greater than 100,000 | 0.82 | (0.55, 1.22) | 0.319 | 0.81 | (0.54, 1.21) | 0.306 |
| Highest level of education (ISCED) | 5: Tertiary | 1 |  |  | 1 |  |  |
|  | 4: Post-secondary non-tertiary | 1.16 | (0.94, 1.43) | 0.169 | 1.16 | (0.94, 1.43) | 0.166 |
|  | 2-3: Secondary | 1.20 | (1.01, 1.43) | 0.043 | 1.20 | (1.01, 1.44) | 0.041 |
|  | 1: Primary | 1.19 | (0.98, 1.45) | 0.081 | 1.20 | (0.99, 1.46) | 0.067 |
| UK country of residence | England | 1 |  |  | 1 |  |  |
|  | Scotland | 0.96 | (0.75, 1.24) | 0.762 | 0.96 | (0.75, 1.24) | 0.771 |
|  | Wales | 0.58 | (0.37, 0.90) | 0.016 | 0.58 | (0.37, 0.90) | 0.015 |

### 5 Checking MR assumptions

#### 5.1 Relevance assumption

Supplementary Table 6: Relevance assumption check for full SBP- and DBP-PRS. Checked via linear regression adjusting for age (continuous), sex, BMI (continuous), smoking status, genetic array and first 10 PCs. F-statistics>10 indicates low risk of weak instrument bias.

|  | Estimate (per mmHg increase in PRS) | Std. Error | t value | P | F-statistic |
| --- | --- | --- | --- | --- | --- |
| **SBP-PRS** | 1.151 | 0.008325 | 138.3 | <0.001 | 6,239 |
| **DBP-PRS** | 1.056 | 0.007803 | 135.4 | <0.001 | 4,634 |

Supplementary Table 7: Relevance assumption check for SBP- and DBP-PRS after removing SNPs associated with confounders (i.e. p< 5e-8).

|  | Estimate (per mmHg increase in PRS) | Std. Error | t value | P | F-statistic |
| --- | --- | --- | --- | --- | --- |
| **SBP-PRS** | 0.5401 | 0.00495 | 109.1 | <0.001 | 4,187 |
| **DBP-PRS** | 1.0647 | 0.00815 | 130.6 | <0.001 | 4,546 |

Supplementary Table 8: Relevance assumption check for SBP- and DBP-PRS after removing SNPs associated with confounders (i.e. p< 5e-6).

|  | Estimate (per mmHg increase in PRS) | Std. Error | t value | P | F-statistic |
| --- | --- | --- | --- | --- | --- |
| **SBP-PRS** | 0.5432 | 0.005024 | 108.1 | <0.001 | 4,173 |
| **DBP-PRS** | 1.0654 | 0.008239 | 129.3 | <0.001 | 4,522 |

#### 5.2 Independence assumption

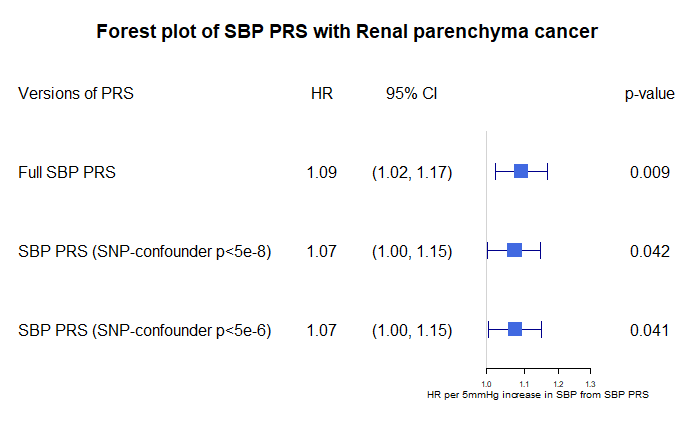

(a) Genetic SBP association with renal-parenchyma cancer

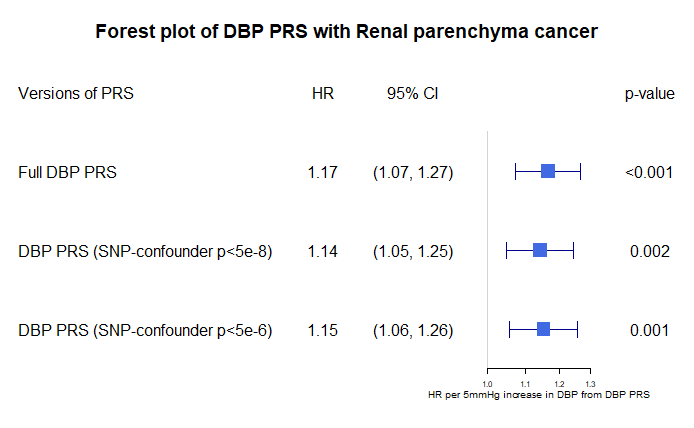

(b) Genetic DBP association with renal-parenchyma cancer

Supplementary Figure 2: Genetic association of (a) SBP conferred by SBP-PRS, and (b) DBP conferred by DBP-PRS, with renal-parenchyma cancer. All models were adjusted for age, sex, BMI, smoking status, genotype array and first 10 genetic PCs. Three versions of PRS were presented: the “Full BP PRS” is the main MR analysis where all SNPs are retained; the remaining two “BP PRS” are sensitivity analyses where pleiotropic variants (associated with BMI, obesity, or smoking status) are removed if their p-value < 5e-8 or 5e-6, respectively.

#### 5.3 Exclusion restriction

##### 5.3.1 Negative control outcomes

Supplementary Table 9: Genetic association of SBP conferred by SBP-PRS with bladder cancer, N= 313,520. “HR” column for SBP presents the HR per 5 mmHg SBP increase from PRS (i.e. $exp\left( \frac{\beta_{ZY}}{\beta_{ZX}}\times5 \right)$). “HR” column of all other covariates is presented as the standard (i.e. HR for increase in one unit of covariate).

| Coefficient | Levels | HR | 95% CI | p |
| --- | --- | --- | --- | --- |
| SBP |  | 0.97 | (0.91, 1.03) | 0.345 |
| Age at recruitment, years |  | 1.11 | (1.10, 1.12) | <0.001 |
| Gender | Female | 1 |  |  |
|  | Male | 3.52 | (3.07, 4.05) | <0.001 |
| BMI, kg/m^2^ |  | 1.02 | (1.01, 1.04) | 0.001 |
| Smoking status | Never | 1 |  |  |
|  | Previous | 1.73 | (1.52, 1.97) | <0.001 |
|  | Current | 2.97 | (2.51, 3.52) | <0.001 |
| Genotype array | Axiom | 1 |  |  |
|  | BiLEVE | 1.20 | (1.01, 1.41) | 0.034 |
| Principal component 1 |  | 1.00 | (0.96, 1.04) | 0.969 |
| Principal component 2 |  | 1.03 | (0.99, 1.07) | 0.148 |
| Principal component 3 |  | 1.02 | (0.98, 1.06) | 0.305 |
| Principal component 4 |  | 0.99 | (0.96, 1.01) | 0.328 |
| Principal component 5 |  | 0.99 | (0.98, 1.01) | 0.313 |
| Principal component 6 |  | 0.99 | (0.96, 1.03) | 0.731 |
| Principal component 7 |  | 1.01 | (0.98, 1.04) | 0.599 |
| Principal component 8 |  | 1.01 | (0.97, 1.04) | 0.706 |
| Principal component 9 |  | 1.01 | (1.00, 1.02) | 0.212 |
| Principal component 10 |  | 1.01 | (0.99, 1.04) | 0.331 |

Supplementary Table 10: Genetic association of DBP conferred by DBP-PRS with bladder cancer, N= 313,520. “HR” column for DBP presents the HR per 5 mmHg DBP increase from PRS (i.e. $exp\left( \frac{\beta_{ZY}}{\beta_{ZX}}\times5 \right)$). “HR” column of all other covariates is presented as the standard (i.e. HR for increase in one unit of covariate).

| Coefficient | Levels | HR | 95% CI | p |
| --- | --- | --- | --- | --- |
| DBP |  | 1.03 | (0.92, 1.16) | 0.611 |
| Age at recruitment, years |  | 1.11 | (1.10, 1.12) | <0.001 |
| Gender | Female | 1 |  |  |
|  | Male | 3.52 | (3.06, 4.05) | <0.001 |
| BMI, kg/m^2^ |  | 1.02 | (1.01, 1.04) | <0.001 |
| Smoking status | Never | 1 |  |  |
|  | Previous | 1.73 | (1.51, 1.97) | <0.001 |
|  | Current | 2.97 | (2.51, 3.52) | <0.001 |
| Genotype array | Axiom | 1 |  |  |
|  | BiLEVE | 1.20 | (1.01, 1.41) | 0.034 |
| Principal component 1 |  | 1.00 | (0.96, 1.04) | 0.958 |
| Principal component 2 |  | 1.03 | (0.99, 1.07) | 0.149 |
| Principal component 3 |  | 1.02 | (0.98, 1.06) | 0.303 |
| Principal component 4 |  | 0.99 | (0.96, 1.01) | 0.327 |
| Principal component 5 |  | 0.99 | (0.98, 1.01) | 0.327 |
| Principal component 6 |  | 0.99 | (0.96, 1.03) | 0.728 |
| Principal component 7 |  | 1.01 | (0.98, 1.04) | 0.596 |
| Principal component 8 |  | 1.01 | (0.97, 1.04) | 0.713 |
| Principal component 9 |  | 1.01 | (0.99, 1.02) | 0.215 |
| Principal component 10 |  | 1.01 | (0.99, 1.04) | 0.331 |

##### 5.3.2 Two-sample MR Analyses

Supplementary Table 11: MR sensitivity analyses for genetically predicted effects of SBP and DBP on renal-parenchyma cancer. SBP: systolic blood pressure. DBP: Diastolic blood pressure. IVW: Inverse variance weighted.

|  | **SBP** | | | **DBP** | | |
| --- | --- | --- | --- | --- | --- | --- |
| **Method** | **Estimate** | **95% CI** | **p** | **Estimate** | **95% CI** | **p** |
| IVW (random effect) | 0.02 | (0.01, 0.04) | 0.009 | 0.05 | (0.02, 0.07) | <0.001 |
| *IVW Heterogeneity test* | NA | NA | 0.548 | NA | NA | 0.594 |
| MR-Egger | 0.03 | (0.00, 0.06) | 0.048 | 0.05 | (0.01, 0.10) | 0.027 |
| *Egger-intercept test* | NA | NA | 0.465 | NA | NA | 0.878 |
| Weighted median | 0.03 | (0.00, 0.05) | 0.032 | 0.05 | (0.00, 0.09) | 0.031 |

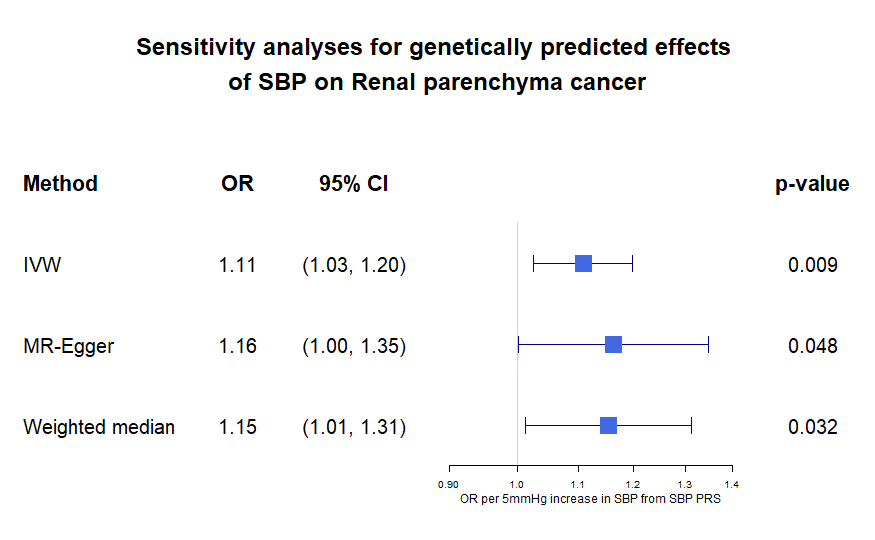

(a)

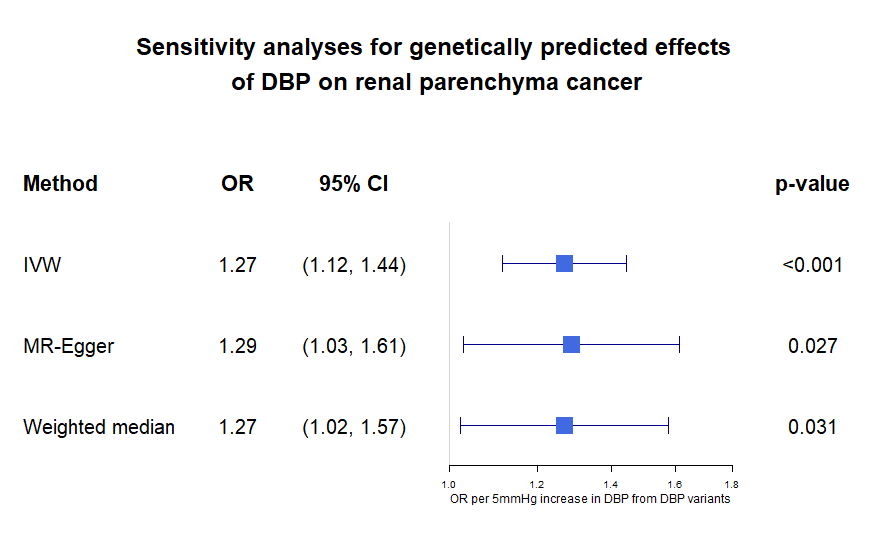

(b)

Supplementary Figure 3: Causal estimates for (a) SBP and (b) DBP obtained from two-sample MR models. Same results as those in Tables 10 and 11 above, but beta estimates were scaled to 5mmHg; p-values are not affected. IVW: Inverse variance weighted.

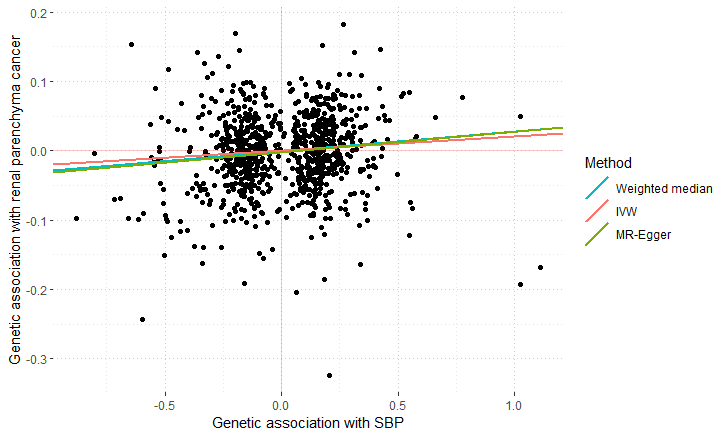

(a)

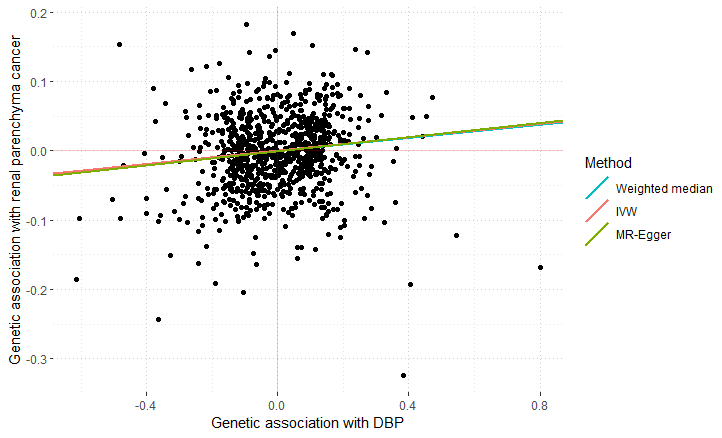

(b)

Supplementary Figure 4: Graphical summary of results obtained from two-sample MR methods (IVW random-effects, MR-Egger, and weighted median methods). Y-axis shows the effect of the SNP on renal parenchyma cancer (i.e. $\beta_{ZY}$). X-axis shows the effect of SNP on exposure (i.e. $\beta_{ZX}$), (a) SBP (b) DBP. Each dot represents an individual SNP. The causal estimate of BP (i.e.$\beta_{\hat{X}Y}$) obtained from each method is displayed as the slope of the line in the corresponding colour. The estimates of the slope correspond to the “Estimate” column in Supplementary Table 11. IVW: Inverse variance weighted. SBP: Systolic blood pressure. DBP: Diastolic blood pressure.

### 6 Removing first 10 PCs

*Supplementary Table 12: Genetic association of SBP conferred by SBP-PRS with renal-parenchyma cancer without adjusting for first 10 PCs, N= 313,520. “HR” column for SBP presents the HR per 5 mmHg SBP increase from PRS (i.e.* $exp\left( \frac{\beta_{ZY}}{\beta_{ZX}}\times5 \right)$*). “HR” column of all other covariates is presented as the standard (i.e. HR for increase in one unit of covariate).*

| Coefficient | Levels | HR | 95% CI | p |
| --- | --- | --- | --- | --- |
| SBP |  | 1.09 | (1.02, 1.16) | 0.009 |
| Age at recruitment, years |  | 1.07 | (1.06, 1.08) | <0.001 |
| Gender | Female | 1 |  |  |
|  | Male | 1.96 | (1.73, 2.21) | <0.001 |
| BMI, kg/m^2^ |  | 1.06 | (1.05, 1.07) | <0.001 |
| Smoking status | Never | 1 |  |  |
|  | Previous | 1.09 | (0.96, 1.24) | 0.187 |
|  | Current | 1.68 | (1.41, 2.00) | <0.001 |
| Genotype array | Axiom | 1 |  |  |
|  | BiLEVE | 1.17 | (0.99, 1.38) | 0.072 |

*Supplementary Table 13: Association of full DBP-PRS with renal-parenchyma cancer without adjusting for first 10 PCs, N= 313,520. “HR” column for DBP presents the HR per 5 mmHg DBP increase from PRS (i.e.* $exp\left( \frac{\beta_{ZY}}{\beta_{ZX}}\times5 \right)$*). “HR” column of all other covariates is presented as the standard (i.e. HR for increase in one unit of covariate).*

| Coefficient | Levels | HR | 95% CI | p |
| --- | --- | --- | --- | --- |
| DBP |  | 1.17 | (1.07, 1.27) | <0.001 |
| Age at recruitment, years |  | 1.07 | (1.06, 1.08) | <0.001 |
| Gender | Female | 1 |  |  |
|  | Male | 1.95 | (1.73, 2.21) | <0.001 |
| BMI, kg/m^2^ |  | 1.06 | (1.05, 1.07) | <0.001 |
| Smoking status | Never | 1 |  |  |
|  | Previous | 1.09 | (0.96, 1.24) | 0.191 |
|  | Current | 1.68 | (1.41, 2.00) | <0.001 |
| Genotype array | Axiom | 1 |  |  |
|  | BiLEVE | 1.17 | (0.99, 1.38) | 0.069 |

### 7 Stratified analysis by sex

Supplementary Table 14: Genetic association of BP conferred by BP-PRS with renal-parenchyma cancer stratified by sex, N= 313,520. “HR” column for BP presents the HR per 5 mmHg BP increase from PRS. SBP: Systolic blood pressure. DBP: Diastolic blood pressure.

|  | Female (N= 166,794) | | | Male (N= 146,726) | | |
| --- | --- | --- | --- | --- | --- | --- |
| Coefficient | HR | 95% CI | p | HR | 95% CI | p |
| SBP | 1.09 | (0.99, 1.21) | 0.088 | 1.10 | (1.00, 1.20) | 0.047 |
| DBP | 1.19 | (0.98, 1.44) | 0.076 | 1.28 | (1.10, 1.50) | 0.001 |

Supplementary Table 15: Genetic association of BP conferred by BP-PRS with renal-composite cancer stratified by sex, N= 313,520. “HR” column for BP presents the HR per 5 mmHg BP increase from PRS. SBP: Systolic blood pressure. DBP: Diastolic blood pressure.

|  | Female (N= 166,794) | | | Male (N= 146,726) | | |
| --- | --- | --- | --- | --- | --- | --- |
| Coefficient | HR | 95% CI | p | HR | 95% CI | p |
| SBP | 1.12 | (1.02, 1.24) | 0.021 | 1.10 | (1.01, 1.20) | 0.030 |
| DBP | 1.23 | (1.02, 1.49) | 0.028 | 1.29 | (1.11, 1.50) | <0.001 |

To further investigate the difference between men and women, we added an interaction term SBP×sex in the above MR model for SBP. Similarly, we added an interaction term DBP×sex in the above MR model for DBP. We did not observe statistically significant interactions for renal-composite cancer (p_SBP×sex_ = 0.60, p_DBP×sex_ = 0.79).

### 8 Results of renal-composite cancer

Supplementary Figure 5 shows that both baseline SBP and DBP were significantly associated with risk of renal-composite cancer, after adjustment for covariates described in the Methods section.

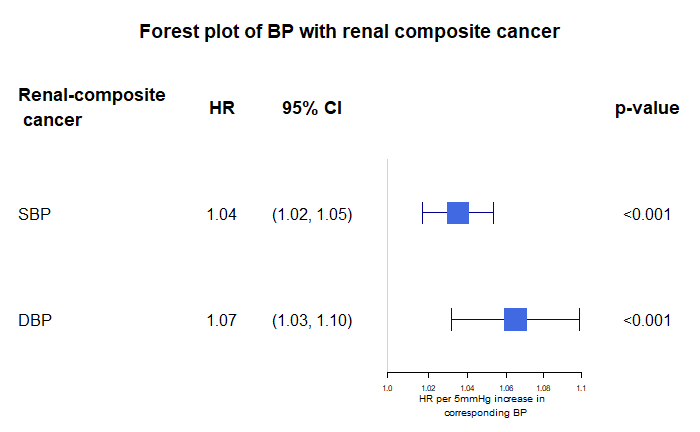

Supplementary Figure 5: Observational association of BP with renal-composite cancer via a multivariable Cox model adjusted for age, sex, BMI, smoking status, weekly alcohol intake, weekly physical activity, the number of comorbidities, Townsend deprivation index, household income, education, and UK country of residence.

Supplementary Figure 6 shows the genetic association of renal-composite cancer obtained from our main one-sample MR analysis. DBP showed stronger effect than SBP on renal-composite cancer, according to the two parallel models described in Methods.

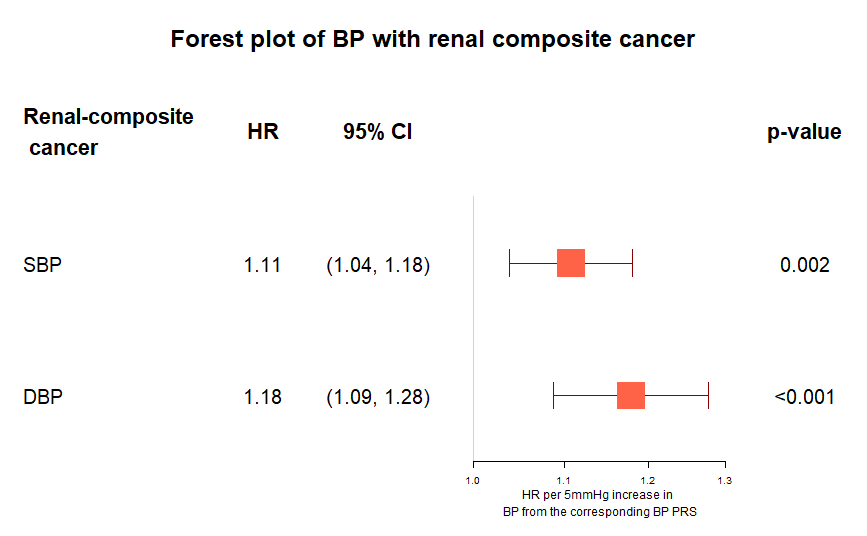

Supplementary Figure 6: Genetic association of BP conferred by the corresponding BP-PRS with renal-composite cancer. All models were adjusted for age, sex, BMI, smoking status, genotype array and first 10 genetic PCs.
